# Microbiome signature of Parkinson disease in healthy and genetically at-risk individuals

**DOI:** 10.1101/2025.05.19.25327907

**Authors:** Elisa Menozzi, Victoria Meslier, Yani Ren, Mallia Geiger, Jane Macnaughtan, Micol Avenali, Marine Gilles, Caterina Galandra, Pierfrancesco Mitrotti, Luca Gallo, Alexandre Famechon, Sara Lucas Del Pozo, Roxana Mezabrovschi, Sofia Koletsi, Nadine Loefflad, Selen Yalkic, Naomi Limbachiya, Hervé Blottière, Christian Morabito, Aymeric David, Benoit Quinquis, Nicolas Pons, Franco Valzania, Francesco Cavallieri, Valentina Fioravanti, Giulia Toschi, Fabio Blandini, Stanislav Dusko Ehrlich, Mathieu Almeida, Anthony HV Schapira

## Abstract

Parkinson disease (PD) is a major cause of disability with significant personal and system financial implications^1,2^. *GBA1* variants are the commonest genetic risk factor for PD and increase the risk up to 30-fold^3,4^. Why only ∼20% of *GBA1* variant carriers develop PD remains unknown. Here, by combining clinical and faecal metagenomics data from PD individuals carrying or not *GBA1* variants, carriers of *GBA1* variants not manifesting PD symptoms (GBA-NMC) and healthy controls, and using an innovative microbiome analysis, we show that a large component of the gut microbiome evolves from healthy across GBA-NMC towards PD and is strongly correlated with disease progression in patients, and prodromal symptoms suggestive of future development of PD in GBA-NMC. By using species altered in PD and GBA-NMC, we detect healthy individuals with the strongest prodromal PD clinical profile. These findings indicate that gut microbiome alterations can identify individuals with *GBA1* variants most prone to develop PD and individuals in the general population that may be progressing towards PD. Healthy nutrition mitigates microbiome alterations associated with disease severity, suggesting that it could modify disease progression in PD patients and decrease the risk of disease development in healthy individuals including those with *GBA1* variants.

## Introduction

Among neurodegenerative disorders, Parkinson disease (PD) is the fastest growing in prevalence, disability, and deaths^1^. The economic burden associated with direct medical costs, disability income, costs for paid care and social productivity loss in the United States is estimated to increase from $52 billion in 2017 to $79 billion in 2037^2^. PD is characterised pathologically by loss of dopaminergic neurons from the substantia nigra pars compacta and accumulation of aggregated α-synuclein in the brainstem and several cortical regions^5^. When motor symptoms appear and a clinical diagnosis is possible, the extent of dopaminergic loss is already greater than 50%^6^. Symptomatic treatment includes dopamine replacement e.g. levodopa^5^. Slowing, halting or preventing the neurodegenerative process before it translates into clinically evident or disabling symptoms requires early detection of those individuals at risk of or progressing towards disease.

The known causes of PD are genetic, including single gene, risk gene and polygenic contributions, possibly combined with as yet unidentified environmental factors. Genetic variants in the *GBA1* gene, encoding the lysosomal enzyme glucocerebrosidase, are found in ∼15% of PD cases^3^, representing the commonest genetic risk factor for PD and one of the most appealing targets for new drug development^3,4^. However, only a proportion of individuals carrying *GBA1* variants will develop PD over their lifetime, with penetrance being estimated at 10% at 60 years up to 19% at 80 years^7^. To date, there are still limited clinical, imaging or biochemical markers that can stratify *GBA1* variant carriers for their future risk of developing PD^8^.

Evidence supports a role of the microbiota-gut-brain axis in the pathogenesis of PD. Although motor symptoms remain the core criteria to diagnose PD, non-motor symptoms such as rapid eye-movement (REM) sleep behaviour disorder (RBD), hyposmia, and autonomic dysfunction including constipation, can predate the onset of motor symptoms by many years in some patients, and thus are referred to as prodromal symptoms^9^. Based on presence or absence of prodromal RBD, PD patients can be classified as body-first or brain-first, with the former predicted to manifest initial signs of neurodegeneration and accumulation of α-synuclein in the autonomic and enteric nervous system with subsequent spread to the central nervous system, while the latter with initial pathology in the brain^10^. Alterations in gut microbiome composition have been detected in individuals with overt PD^11,12^, especially in body-first PD^13^. Moreover, gut dysbiosis can precipitate PD in animal models^14^, and exposure to microbial components can induce α-synuclein accumulation in gut enteroendocrine cells^15^.

By comparing gut microbiomes of a cohort of PD patients (n=271), disease-free *GBA1* variant carriers (n=43) and healthy controls (n=150), we find large scale PD-specific alterations in individuals of the last two groups. These alterations are associated with a PD prodromal clinical profile and are modulated by a healthy diet. We suggest that alterations in gut microbiome composition might explain the incomplete penetrance of PD in *GBA1* variant carriers, and help identify those at highest risk of conversion to PD. We further suggest that similar changes in healthy individuals without known genetic risk may help identify those predisposed towards PD development and that healthy diet might mitigate this development.

## Results

### Study participants

A total of 540 participants were included in the full analysis set for clinical data. Participants’ characteristics are shown in Table 1. The PD participants (n=314) included both carriers (n=128) and non-carriers (n=186) of *GBA1* variants. The individuals without PD included healthy controls non-carriers of *GBA1* variants (n=175, HC) and non-manifesting *GBA1* variant carriers (n=51, GBA-NMC). When demographic features of GBA-NMC were compared to the other groups, no differences were detected. PD participants were significantly older than HC (p=0.006), and more frequently males (p=0.028). More than half of the HC participants were partners of people with PD, to mitigate the effect of diet or other lifestyle associated variables on gut microbiome composition.

**Table 1.** Overview of study cohort. BMI, body mass index; BDI, Beck Depression Inventory; DBS, deep brain stimulation; DQS, Dietary Quality Score; HADS, Hospital Anxiety and Depression Scale; H&Y, Hoehn & Yahr ; LCIG, levodopa-carbidopa intestinal gel; LEDD, levodopa equivalent daily dose; GBR, Great Britain; ITA, Italy; MDS-UPDRS, Movement Disorder Society (MDS) Unified Parkinson’s Disease Rating Scale; MOCA, Montreal Cognitive Assessment; PD, Parkinson disease; SCOPA-AUT, SCales for Outcomes in PArkinson’s disease; RBDSQ, REM Sleep Behavior Disorder Questionnaire; UPSIT, University of Pennsylvania Smell Identification Test; WCSS, Wexner Constipation Scoring System. Significance of P values: ns (not significant): p ≥ 0.05; *, p<0.05; **, p<0.01; ***, p<0.001; **** p<0.0001.

|  | HC<br>(N=175) | GBA-NMC<br>(N=51) | PD<br>(N = 314) | *p* (HC vs<br>GBA-NMC) | *p* (GBA-NMC vs<br>PD) |
| --- | --- | --- | --- | --- | --- |
| **DEMOGRAPHICS** |  |  |  |  |  |
| **GBR cohort (N)** | 87 | 40 | 155 | - | - |
| **ITA cohort (N)** | 88 | 11 | 159 | - | - |
| **Age** | 60 ± 10 | 61 ± 11 | 63 ± 9 | ns | ns |
| **Sex (% F, N)** | 58% (102) | 57% (29) | 44% (139) | ns | ns |
| **Positive PD Family History (% , N)** | 32% (56) | 51% (26) | 29% (90) | * | ** |
| **Education (yrs)** | 14.7 ± 4.3 | 15.8 ± 4.0 | 14 ± 4.3 | ns | * |
| **Participation with partner (% , N)** | 52% (91) | 27% (14) | 26% (83) | ** | ns |
| **Participation with family member (% , N)** | 12% (21) | 41% (21) | 10% (32) | **** | **** |
| **BMI (kg/m<sup>2</sup>)** | 25.6 ± 4.4 | 26.1 ± 3.9 | 25.7 ± 4.9 | ns | ns |
| **DISEASE-ASSOCIATED FEATURES** |  |  |  |  |  |
| **PD Age at onset** | - | - | 57 ± 10 | - | - |
| **PD Duration** | - | - | 6.4 ± 5 | - | - |
| **Drug naïve (% , N)** | - | - | 11% (36) | - | - |
| **LEDD (mg)** | - | - | 590.7 ± 452.8 | - | - |
| **DBS (% , N)** | - | - | 13% (40) | - | - |
| **Apomorphine pump infusion (% , N)** | - | - | 0.3% (1) | - | - |
| **LCIG (% , N)** | - | - | 0.3% (1) | - | - |
| **CLINICAL FEATURES** |  |  |  |  |  |
| **MDS-UPDRS part I** | 3.4 ± 3.9 | 5.2 ± 4.7 | 10.3 ± 6.6 | ** | **** |
| **MDS-UPDRS part II** | 0.4 ± 1 | 1.2 ± 1.8 | 10.8 ± 6.6 | *** | **** |
| **MDS-UPDRS part III** | 2.2 ± 3.3 | 3.5 ± 3.8 | 27.4 ± 12.9 | * | **** |
| **MDS-UPDRS part IV** | - | - | 3.1 ± 3.5 | - | - |
| **MDS-UPDRS total** | 6 ± 6.4 | 9.9 ± 7.5 | 51.3 ± 22.1 | *** | **** |
| **H&Y stage (median)** | 0 | 0 | 2 | ns | **** |
| **SCOPA-AUT total** | 7.2 ± 5.1 | 7.9 ± 5.4 | 14.4 ± 7.8 | ns | **** |
| **SCOPA Gastrointestinal** | 1.1 ± 1.3 | 1.2 ± 1.3 | 3.9 ± 2.9 | ns | **** |
| **SCOPA Urinary** | 2.8 ± 2.3 | 3.7 ± 2.7 | 4.9 ± 3.1 | * | ** |
| **SCOPA Cardiovascular** | 0.2 ± 0.6 | 0.4 ± 0.7 | 0.8 ± 1.1 | ns | ** |
| **SCOPA Thermoregulatory** | 1.2 ± 1.4 | 1.3 ± 1.7 | 2.2 ± 2.2 | ns | ** |
| **SCOPA Pupillomotor** | 0.4 ± 0.7 | 0.5 ± 0.7 | 0.5 ± 0.8 | ns | ns |
| **SCOPA Sexual** | 0.9 ± 1.2 | 0.7 ± 1.2 | 1.5 ± 1.8 | ns | ** |
| **WCSS** | 2.5 ± 2.5 | 3.2 ± 3.2 | 5.9 ± 4.5 | ns | **** |
| **RBDSQ (raw score)** | 1.6 ± 2 | 2.3 ± 2.3 | 5.3 ± 3.6 | * | **** |
| **RBD above cut-off (%**<br>**N)** | 10% (17) | 18% (9) | 39% (122) | ns | ** |
| **UPSIT** | 29.6 ± 4.5 | 29.7 ±<br>4.8 | 18.3 ± 6.3 | ns | **** |
| **HADS anxiety** | 4 ± 3.3 | 4.1 ± 3.5 | 5.7 ± 3.8 | ns | ** |
| **HADS depression** | 2.7 ± 2.9 | 2.7 ± 2.7 | 5.4 ± 3.7 | ns | **** |
| **BDI** | 5.2 ± 5.4 | 6 ± 6.7 | 9.8 ± 6.7 | ns | **** |
| **MOCA** | 27.3 ± 2.1 | 27 ± 2.9 | 25.7 ± 3.8 | ns | ns |
| **MOCA below cut-off**<br>**(%, N)** | 18% (32) | 29% (15) | 38% (117) | * | ns |

### Clinical profile of GBA-NMC

To identify possible clinical elements that could stratify GBA-NMC for their risk of developing PD, we compared the severity of motor and non-motor symptoms between HC and GBA-NMC, using a wide range of clinical scales and questionnaires (Table 1). We found worse motor symptoms in the GBA-NMC group, either subjectively reported (MDS-UPDRS part II, p = 0.0008), or objectively assessed (MDS-UPDRS part III, p = 0.015). In terms of non-motor symptoms, no differences in constipation or global autonomic function were detected, however GBA-NMC showed higher scores in the urinary sub-score of the SCOPA-AUT (p = 0.017) and in the MDS-UPDRS part I (p=0.004). GBA-NMC were more likely to show cognitive impairment compared to HC (β = 0.88, p = 0.02, OR 2.4, 95% CI 1.2-5.3). No differences in depression, anxiety, or olfactory function were observed. Within the GBA-NMC, 10 individuals reached the threshold for estimated probability according to the MDS prodromal criteria calculated based on available information (listed in Supplementary Methods)^16^.

Overall, our clinical data suggest that in our GBA-NMC cohort, there may be a group of individuals who exhibit some prodromal symptoms (e.g., subthreshold parkinsonism, some dysautonomia), and thus might be in their prodromal phase of PD.

### Significant microbiome alterations in PD

Microbiome profiles were successfully generated for 464 individuals (150 HC, 43 GBA-NMC and 271 PD). We first compared microbiomes of PD patients, carriers (n=109) or non-carriers (n=162) of *GBA1* variants, and found 44 Metagenomic Species Pangenomes (MSPs) out of 627 with prevalence of at least 10% in our cohort that were different in abundance at p<0.05 by Wilcoxon rank sum test. However, none remained significantly different after Benjamini-Hochberg correction for multiple testing at q<0.1 (or even q<0.6). In contrast, the comparison of either group with healthy controls (HC, n=150) revealed numerous MSPs significant at q<0.1 (82 and 123 for carrier and non-carrier group, respectively; the difference is likely due to the higher statistical power for the more numerous non-carrier group). We conclude that, in overt PD, *GBA1* genetic status has little or no impact on the microbiome composition, whereas the disease itself has a strong impact.

We therefore pooled all PD patients regardless of their genetic status (n=271), to identify the PD gut microbiome signature at maximal statistical power and compared them with HC (n=150). A total of 176 species were differentially abundant (p<0.05, Supplementary Table 1, Extended Fig. 1A). In PD, Actinobacteriota were enriched at phylum level and *Bifidobacteriaceae* at family level (Supplementary Table 2), as previously reported^12,17,18^. Three other families of Firmicutes, *Streptococcaceae*, *Lactobacillaceae* and *Lachnospiraceae J*, were enriched to significance by Chi^2^ test (p<0.05); interestingly, the former two include oral species. In contrast, species enriched in HC belonged to *Lachnospiraceae C*, *Oscillospiraceae*, and *Lachnospiraceae K*. These families include butyrate producers such as *Roseburia* and *Dysmobacter*, which may be anti-inflammatory. *Bifidobacterium* and *Streptococcus* genera were enriched in PD, while *Dysmobacter-Oscillibacter* and *Faecalibacterium* were enriched in HC. Species that showed the greatest increase in PD included *Streptococcus mutans*, *Bifidobacterium longum* and *B. dentium* and *Lactobacillus paragasseri*, whereas species that showed the greatest depletion in PD included *Roseburia intestinalis* and *R. inulinivorans* and an unclassified *Faecalibacterium*.

### An intermediate and consistent microbiome alteration in GBA-NMC

We then investigated whether some of the PD-related gut microbiome alterations were detectable in the GBA-NMC group (n=43) relative to HC (n=150).

We found 43 species with significantly different abundance between HC and GBA-NMC (Supplementary Table 3, Extended Fig. 1B), less than between HC and PD, possibly, at least in part, because of the loss of statistical power due to the low number of GBA-NMC. Of these, 21 were common with the 176 species significantly altered in PD patients compared to HC, and all but one were enriched or depleted in GBA-NMC individuals coherently with enrichment or depletion in PD patients, which is unlikely to happen by chance (Chi^2^ test, p=5.7E-3). The effect sizes observed for the common MSPs in the comparisons between GBA-NMC vs HC, and PD vs HC were highly correlated (red dots, Fig. 1A).

**Fig. 1:**
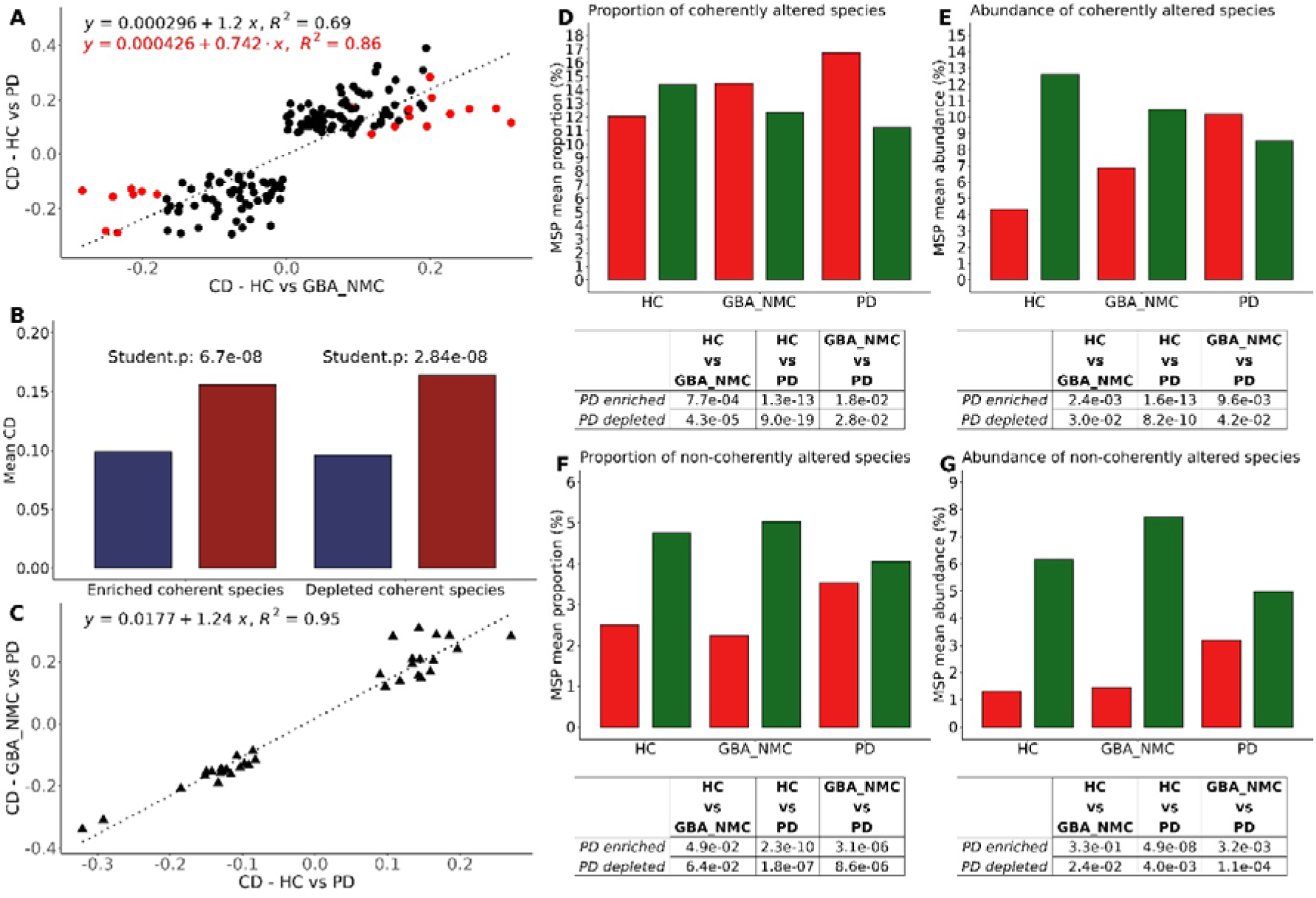
Variation of the coherent and non-coherent gut microbiome species in HC individuals, GBA-NMC participants and PD patients. A. Correlations of Cliff Deltas (CD) of 142 coherent species found by comparing HC individuals with GBA-NMC or PD patients. Negative CD values are species found enriched in HC while positive CD values are species found enriched in both GBA-NMC and PD. Red dots highlight the 20 significant and coherently altered species (Wilcoxon p<0.05) overlapping between HC-PD and HC-GBA-NMC comparisons. R^2^ denotes squared Pearson correlation coefficient when considering the 142 or 20 species. B. Average CD of coherently enriched (n=81) or depleted (n = 61) species in GBA-NMC (blue) and PD (red) relative to HC. C. Correlation of CDs of 34 species altered significantly (Wilcoxon p<0.05) and non-coherently in the comparison of HC and PD patients and in the comparison of PD and GBA-NMC. D and E. Abundance and proportion of coherently altered species in HC, GBA-NMC and PD and corresponding p values (determined by the Student t test for different comparisons); red and green refer to coherently enriched and depleted species, respectively. F and G. Abundance and proportion of non-coherently altered species in the three study groups and corresponding p values; red and green refer to non-coherently enriched and depleted species, respectively.

Importantly, examination of the coherence of variation of the 176 MSPs significantly different between HC and PD individuals, using the direction of Cliff’s delta (CD) as a guide, revealed that 142 were coherently altered in GBA-NMC and PD relative to HC (Supplementary Table 4), which is very unlikely to happen by chance (Chi^2^ test, p=3.9E-16). Effect sizes of these species in GBA-NMC and PD relative to HC were highly correlated (Fig. 1A), and were, on average, significantly lower in GBA-NMC than in PD (Fig. 1B), suggesting that the GBA-NMC microbiome may be in an intermediate state between that of HC and PD. The remaining 34 species were found not to vary coherently in GBA-NMC and PD relative to HC and are termed non-coherent species hereafter; 18 were enriched and 16 depleted in PD (Supplementary Table 4). Their CD in GBA-NMC and HC relative to PD were highly correlated (Fig. 1C). Among the coherent species, we observed enrichment of oral residents (*S. mutans* and *L. paragasseri*) and pro-inflammatory *R. gnavus* and the depletion of butyrate producers (*Roseburia* or *F. prausnitzii*). In contrast, non-coherent enriched species included *Bifidobacteria*, one of the characteristic alterations of the PD gut microbiome^17^, suggesting that their enrichment may take place at the clinical onset of the disease or develop during its course.

The species coherently altered in GBA-NMC and PD relative to HC represented a similar fraction (slightly over 25%) of the microbiome in all study groups, but the abundance and proportion of PD-enriched species significantly increased from HC over GBA-NMC to PD, while the abundance and proportion of species PD-depleted significantly decreased (Fig. 1D-E). The non-coherent species represented about 7-9% of microbiome abundance and 7% of species proportion. This part of the microbiome varied little between HC and GBA-NMC, but very significantly increased and decreased for enriched and depleted species, respectively, in PD relative to HC or GBA-NMC (Fig. 1F-G), supporting the view that it evolves mostly once PD is manifest.

Summarizing, over a quarter of the gut microbiome significantly changes in PD relative to HC, of which we distinguish two components. A major one evolves consistently from HC across GBA-NMC to PD, the extent of changes being lower in GBA-NMC than PD. A minor component changes abruptly in overt PD. Since similar coherent changes in gut microbiome composition are observed in at-risk individuals, such as GBA-NMC, and PD patients, we suggest that the coherently altered species of the PD gut microbiome may represent a prodromal feature of PD (the “prodromal-PD microbiome”), possibly contributing to PD development.

### Associations of microbial alterations and clinical variables in PD

To investigate whether changes in gut microbiome observed in PD patients were associated with specific clinical features, we compared clinical variables in patients belonging to quartiles with highest (n=68) and lowest (n=68) microbiome alterations (Extended Fig. 2A-D). We carried out 8 comparisons based on the type of measure (abundance/proportion) and species (coherent/non-coherent, enriched/depleted). Extended Table 1 shows the results from the comparison of the abundance of coherent depleted species; all comparisons are displayed in Supplementary Table 5. When p values were significant in 3 comparisons or less, we considered them to be only weakly or even not correlated with microbiome alterations. Among these, we found age, BMI and sex, indicating that they do not affect greatly our analyses. When the clinical parameters recurred in at least 6 different comparisons we suggest that these parameters were highly correlated with microbiome alterations (in bold in Extended Table 1; Fig. 2). The clinical parameters highly correlated with microbiome alterations fell in two classes.

**Fig. 2:**
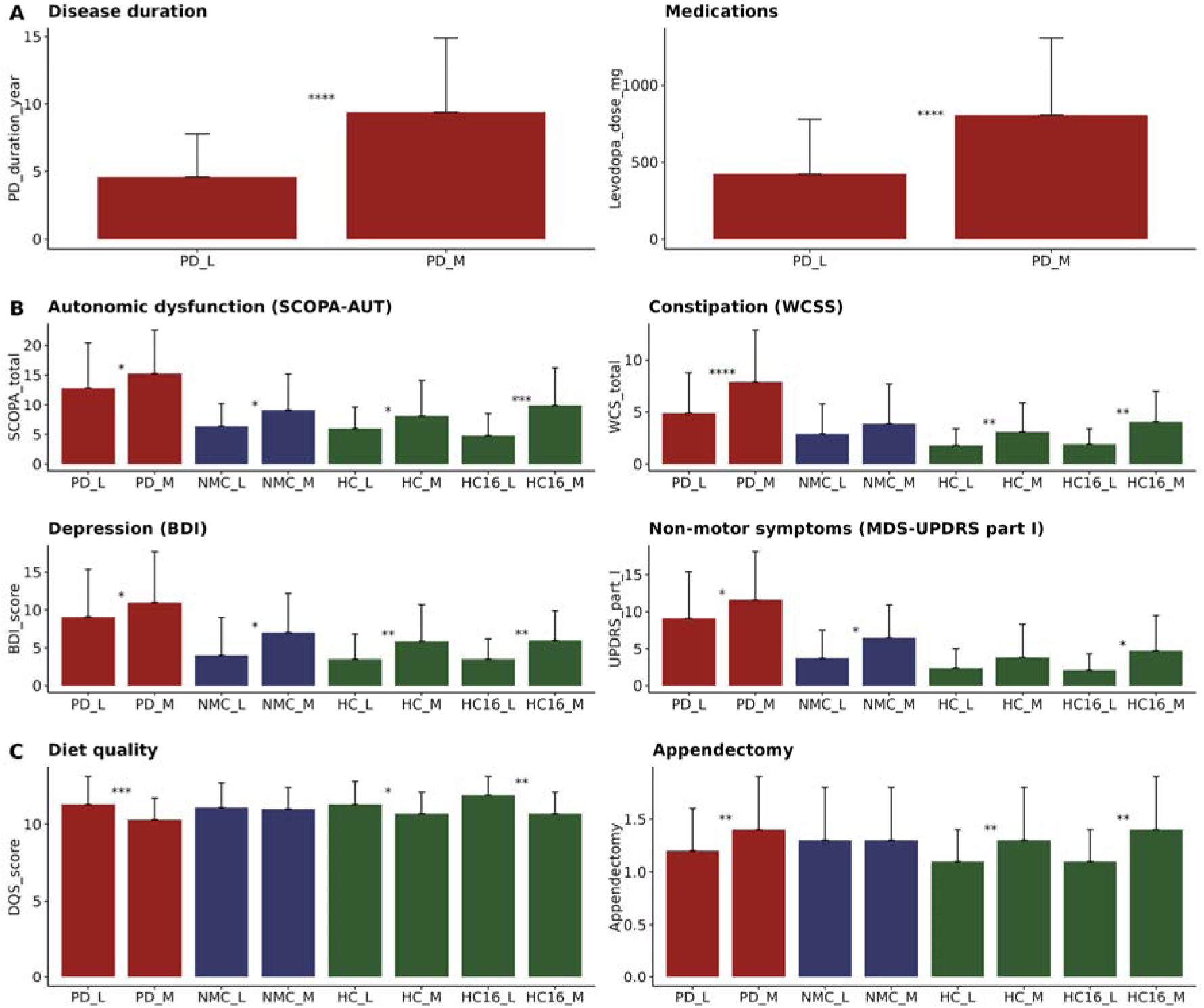
Association of microbiome alterations with clinical variables. Comparison of individuals with least (L) and more (M) altered microbiome across the study groups: for PD individuals, top and bottom quartiles of the distribution according to the abundance of coherent depleted species (n=68 each); for GBA-NMC individuals, those below and above the median according to the proportion of coherent enriched species (n=21 and 22 respectively); for HC individuals, top and bottom quartiles of the distribution according to the proportion of coherent enriched species (HC_L and HC_M, n=38 each). HC individuals having PDMS-16 ≤ -5 (HC16_L, n=21) and PDMS16 ≥3 (HC16_M, n=27) are also shown. A. Disease-associated variables of PD patients (disease duration, and medication dosage); B. Clinical variables for all groups (automatic dysfunction, constipation, depression, non-motor symptoms); C. Health related variables for all groups (diet quality and appendectomy encoded as 1 if individuals did not undergo appendectomy and 2 if people underwent appendectomy). The values of clinical variables are the means listed in Extended Table 1, standard deviations and statistical significance are indicated by the thin bars and stars indicate the significance level of the Student t test; **** for p<0.0001, *** for p< 0.001, ** for p<0.01 and * for p<0.05.

The first class corresponded to variables associated with disease severity, found to be higher in individuals in the top quartile of microbiome alterations. Among these, we detected depression, autonomic dysfunction, constipation and motor dysfunction. A higher percentage of individuals who underwent appendectomy prior to PD development was found in the top quartile; association of appendectomy and PD has been reported but remains controversial^19–23^.

The second class included variables associated with health, found to be worse in individuals with more altered microbiome. These variables included cognitive and olfactory functions, and variables related to nutrition, such as the dietary quality score (DQS) or fruit and vegetable consumption, suggesting a possible relation between food quality and milder disease profile.

We noted that the two most significantly differing variables between top and bottom quartiles were PD duration and levodopa equivalent daily dose (i.e., the amount of anti-parkinsonian medications), both associated with disease severity (Extended Table 1). We thus compared the microbiome of PD patients medicated or drug naïve, adjusting for disease duration and the microbiome of PD patients with different disease duration, adjusting for medication dose (Extended Fig. 3A-B). There were no significant differences in the level of PD-enriched or depleted species in medicated and non-medicated patients with the same disease duration whereas there were significant differences in patients with the same medication dose and different disease duration. We suggest that microbiome evolves with the progression of the disease rather than in response to treatment.

### Associations of microbial alterations and clinical variables in disease-free GBA-NMC and HC

To investigate whether the PD microbiome signature was associated with any prodromal clinical profile in GBA-NMC, we compared clinical variables of GBA-NMC individuals with lower and higher microbiome alterations, below or above the median of the abundance and proportion of coherently PD-enriched/depleted species (Extended Fig. 4; split around median rather than comparison of quartiles was used to conserve statistical power). We identified numerous significantly different parameters, mainly representative of non-motor symptoms which can be present in the prodromal phase (Fig. 2, Extended Table 1, Supplementary Table 6). Among these, depression, anxiety and autonomic dysfunction, were all significantly stronger in individuals showing microbiome alterations above the median. Importantly, all GBA-NMC individuals who were identified as prodromal PD, based on the MDS research criteria^16^, had the abundance of coherently PD-enriched species above the median, a highly significant bias (Chi^2^ test, p=4.7E-3) and 4 were at the very top of the distribution (Fig. 3).

**Fig. 3:**
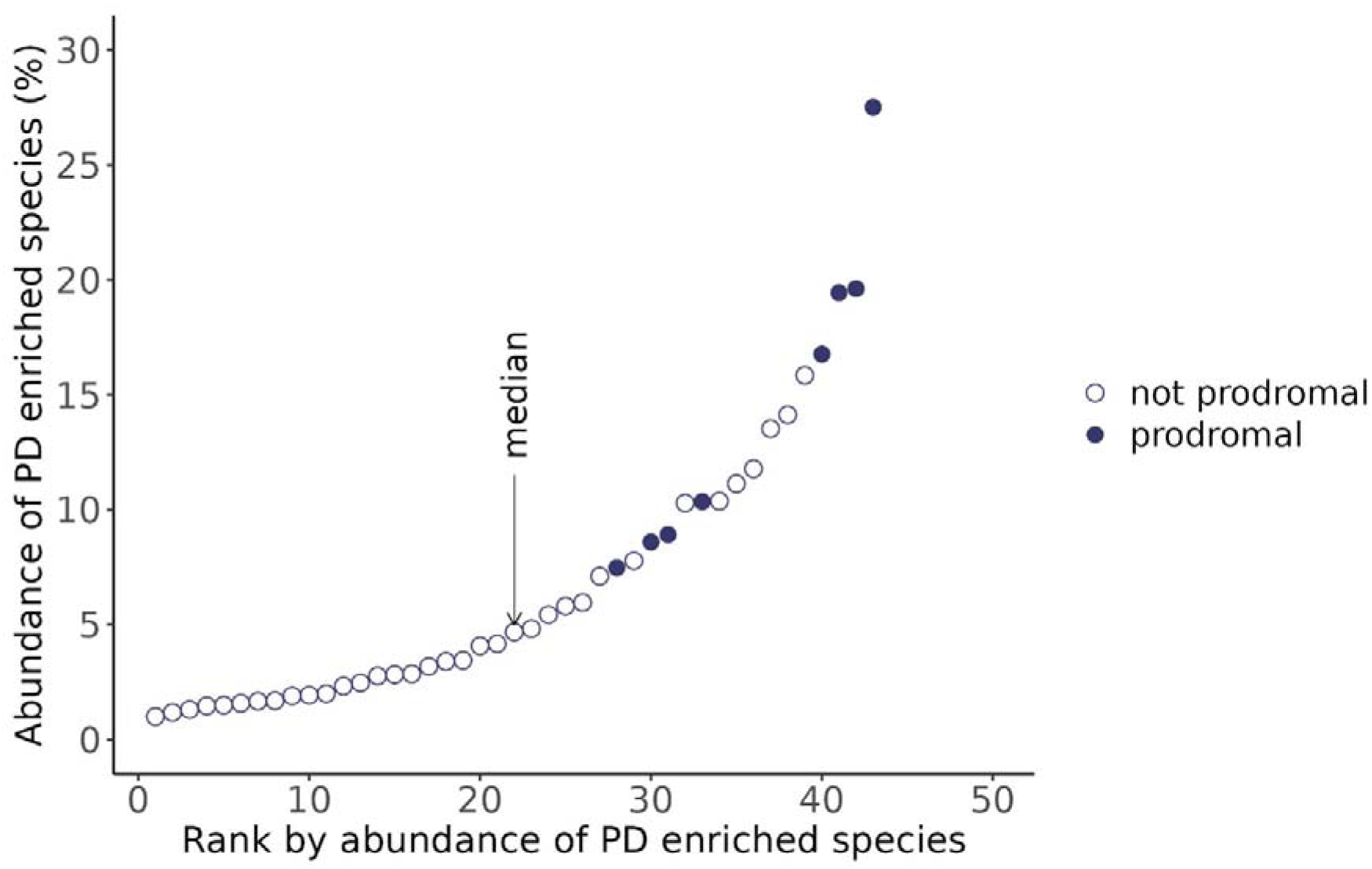
GBA-NMC individuals (n=43) ranked by the abundance of PD enriched species. Individuals assigned to the prodromal group by the MDS research criteria (n=8) are represented in filled dots.

We conclude that GBA-NMC individuals presenting with the microbiome signature closer to PD are the ones showing the strongest prodromal profile, especially in terms of autonomic and peripheral nervous system dysfunction (body-first PD). Paralleling the observation with PD patients, where the duration of overt disease appears to be strongly associated with microbiome alterations, we suggest that in the GBA-NMC group the progression toward overt PD is associated with progressive microbiome alterations and that a combination of clinical and microbiome assessments may help identify those individuals that will convert to disease.

We then investigated microbiome alterations across quartiles of distribution in healthy individuals and found very significant differences, resembling those observed for PD patients (Extended Fig. 5). Interestingly and importantly, comparison of clinical variables of HC individuals assigned to the extreme quartiles, denoted as lower (n=38) and higher (n=38) microbiome alterations for analogy with the PD and GBA-NMC groups (Fig. 2, Extended Table 1 and Supplementary Table 7), revealed significant differences of most variables differing in the GBA-NMC group, including depression, anxiety and autonomic dysfunction. Furthermore, these HC differed in eating habits (DQS, fruit and vegetable consumption), and appendectomy frequency, as observed for PD patients. We conclude that microbiome alterations resembling those of PD patients take place in a healthy population free of known genetic risk, raising the possibility that, as in the individuals with a known genetic risk, such alterations might be associated with progression towards overt disease.

### Correlation of microbial species and clinical variables

We next investigated whether the species that compose the altered microbiome might individually be correlated with clinical variables, focusing on the 176 species differentially abundant in HC and PD. In the PD group, abundance variations of 95% of species were significantly correlated with at least one clinical variable (Extended Fig. 6A, Supplementary Table 8); the number of species correlated with a variable is shown in Fig. 4A. PD-enriched and PD-depleted species were associated positively and negatively, respectively, with disease severity (e.g., non-motor symptoms such as constipation, autonomic dysfunction, depression, RBD) and conversely, negatively and positively with health (cognition and olfactory function; the DQS score).

**Fig. 4:**
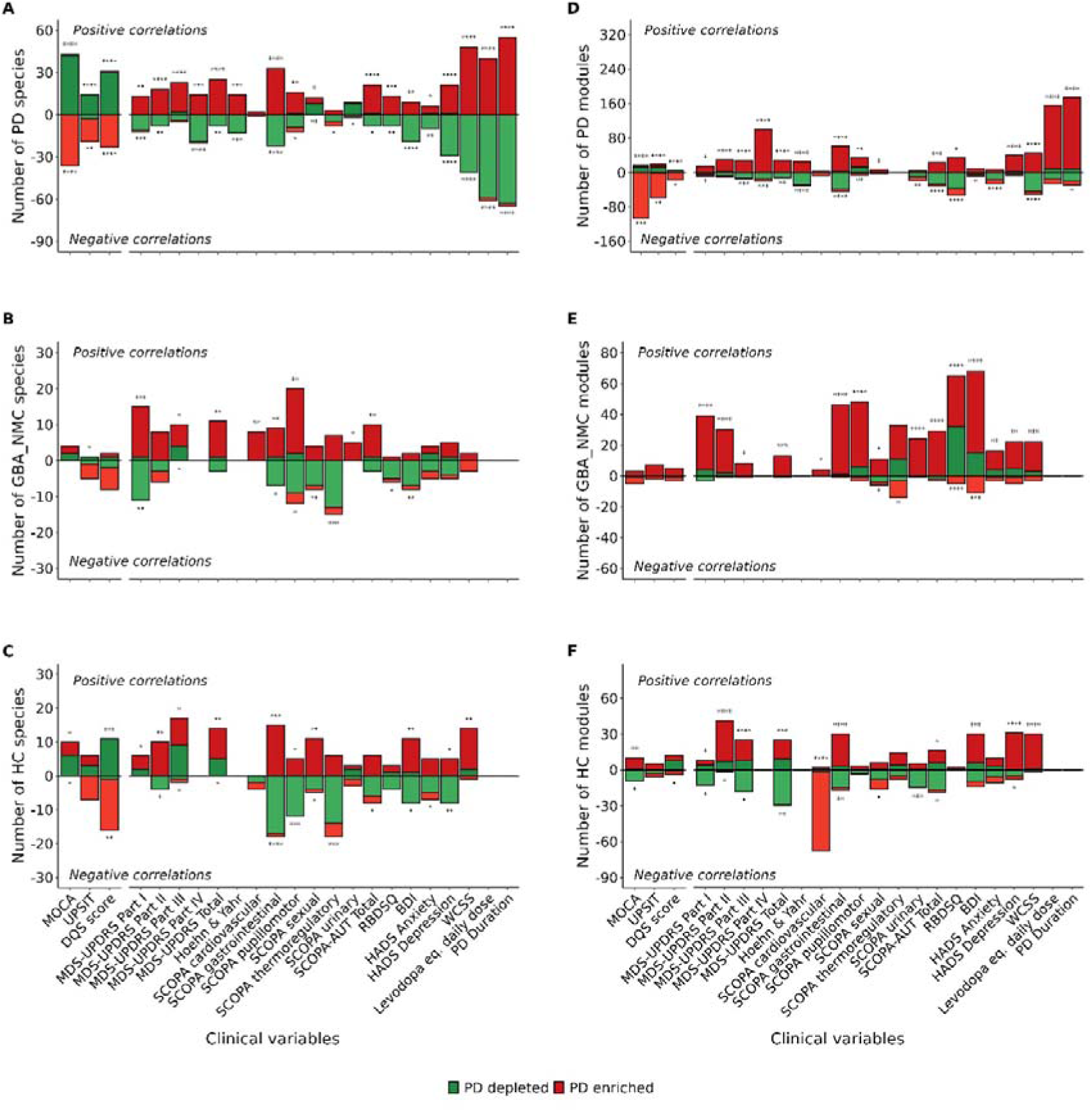
Correlation of microbial species and functional modules with clinical parameters. A to C: Number of microbial species significantly correlated with different clinical parameters; Spearman correlation coefficient with p<0.05 is indicated on the ordinate for PD patients (A), GBA-NMC participants (B) and HC individuals (C). Red and green colors refer to species enriched and depleted in PD, positive and negative integers to positive and negative correlations, respectively. D to F: Number of functional modules correlated significantly with different clinical parameters; Spearman correlation coefficient with p<0.05 is indicated on the ordinate for PD patients (D), GBA-NMC participants (E) and HC individuals (F). Red and green colors refer to modules enriched and depleted in PD, positive and negative integers to positive and negative correlations, respectively. Stars indicate the significance of the correlation bias of enriched and depleted species (Chi2), on the positive and negative half of the graph, respectively; **** for p<0.0001, *** for p< 0.001, ** for p<0.01 and * for p<0.05.

Importantly, we found a similar profile of correlations between species and clinical variables in GBA-NMC (Fig. 4B), the variables associated with severity being mostly positively correlated with species enriched in PD and negatively with species depleted in PD (Extended Fig. 6B); the converse was observed for variables associated with health. The correlations involved 64% of the 176 species (Supplementary Table 9). Some 15% of correlations were identical with those in PD patients (Supplementary Table 10) and an overwhelming majority of those (40/43) were coherent in both groups (Extended Fig. 7A). Other correlations were different, possibly due to relatively small variations of species abundances within a study group, resulting in a statistical significance in one group but not another. Furthermore, similar correlations were found in healthy individuals (Fig. 4C, Supplementary Table 11). They involved 78% of the 176 species, 24% of the correlations were identical to those in the PD group (Supplementary Table 12) and were overwhelmingly coherent with PD (75/76, Extended Fig. 7B).

A striking conclusion from our analyses is that the microbial species correlated with disease severity in PD patients appear correlated with clinical variables that estimate disease severity both in healthy individuals at higher risk of PD development due to their *GBA1* genetic background, and in those without known genetic predisposition.

### Stratification of HC individuals using microbiome features

In view of similar correlations between clinical variables and microbial species in all study groups, we explored the possibility of stratifying HC individuals that may be closer to disease development by the microbial species they harbor. To do so, we first selected the species involved in the same significant correlations with clinical variables in PD and the disease-free GBA-NMC (33 species). Among those, we then selected species with the highest prevalence difference in PD relative to HC (over 10%) and the species correlated coherently with PD duration in patients. As a result, we identified 16 species, of which 10 were enriched and 6 depleted in PD (Supplementary Table 13). By summing the number of PD-enriched species and deducing the number of PD-depleted species that each individual harbored we computed a score, termed PD Microbiome Score-16 (PDMS-16), referring to the number of species used. PDMS-16 distribution among HC and PD individuals is shown in Extended Fig. 8.

We then compared clinical variables between the subgroup of HC presenting with the highest PDMS-16 (3 to 7; n=27, 18% of the group) with i/ 21 individuals with the lowest PDMS-16 score (-5 and -6) and ii/ the remainder of the HC group (n=123). The individuals with the highest PDMS-16 score had more severe depression and anxiety, autonomic dysfunction, and constipation, consumed less healthy food, and had a more frequent history of appendectomy (Fig. 2, Extended Table 1). We conclude that PDMS-16 score can identify individuals with clinical profile closer to that of PD patients than the remainder of the HC group.

### Alterations of microbiome functions in PD and GBA-NMC

Comparison of HC with PD patients revealed that 180 of the 357 complete functional microbial modules from 3 sources (Kyoto Encyclopedia of Genes and Genomes or KEGG, Gut Metabolic Modules or GMM, Gut Brain Modules or GBM), were differentially abundant: 143 were enriched in PD and 37 were depleted (Supplementary Table 14). There was no significant bias of module database (KEGG n=94, GMM n=65, GBM n=21; Chi^2^ test p=0.54).

The PD microbiome was enriched for modules related to neurotransmitters metabolism (dopamine, acetylcholine and GABA), nucleic acid degradation, and amino acid degradation (threonine, phenylalanine, putrescine). Enrichment of the modules for dopamine degradation and DOPAC synthesis (MGB023 and MGB024, respectively) could possibly be due to levodopa treatment of PD patients, as Actinobacteria reportedly can degrade and utilize dopamine^18^. The degradation pathway of the bacterial by-product of protein degradation, p-cresol, was also found to be enriched in our PD cohort. The modules depleted in PD were involved in amino acid synthesis (serine, tryptophan, lysine, tyrosine), riboflavin biosynthesis, dietary carbohydrate degradation (lactose, mannose) pathways and the conversion of acetyl-coenzyme A into acetate pathway. These findings, combined with the enrichment in pathways implicated in the conversion of acetate into acetyl-coenzyme A, or of propionate into succinate, and the tricarboxylic acid (TCA) cycle, support a metabolic shift from carbohydrate fermentation towards proteolysis as an energy source, in line with previous findings^17,24^.

Comparison of HC and GBA-NMC individuals identified only 5 significantly different modules. This low number may reflect, in part, less alterations of functions in the GBA-NMC group, and in part, insufficient statistical power. We therefore examined the coherence of CD variation of the modules significantly different in HC and PD in the GBA-NMC group. Of the 180 modules, 143 varied coherently and 37 non-coherently (Supplementary Table 14), a very significant bias (Chi^2^ test, p=2.8E-15). CDs of the coherently varying modules were significantly correlated (Extended Fig. 9A), and significantly higher in the HC vs PD comparison than in the HC vs GBA-NMC comparison (Extended Fig. 9B). We conclude that functional potential of the microbiome displays variations like those observed for the taxonomic differences.

### Correlations of microbiome functions and clinical variables

Abundance variations of 92% of modules were significantly correlated with at least one of 25 clinical variables in the PD group (Supplementary Table 15). Correlations were biased in a similar way observed for individual species: enriched modules largely correlated positively with variables related to the severity of the disease and negatively to those related to health, while the depleted modules had an opposite bias (Fig. 4D). Similar correlation profiles were observed for the other two study groups, GBA-NMC and HC (Fig. 4E-F, Supplementary Tables 16 and 17), albeit with lower number of modules, likely due to lower statistical power (58.3% and 72.5% for GBA-NMC and HC, respectively). We conclude that, expectedly, functional potential of the microbiome correlates with clinical variables similarly to species that encode it.

## Discussion

Using combined clinical and metagenomics data from a large population of individuals with overt PD and those harbouring genetic predisposition to PD (GBA-NMC), we identify similar, coherent alterations in the gut microbiome profile of PD patients, genetically at-risk individuals presenting a higher burden of prodromal symptoms and in approximately 20% of healthy individuals without known genetic predisposition for PD. This was possible by the examination of the coherence of microbiome alterations between the study groups, in addition to the more common assessment of their statistical significance, allowing detection of small abundance alterations of many species, and not only large alterations of a small number of species. This innovative paradigm of microbiome analysis may be useful in studies with a design like ours, where at-risk groups are compared with both healthy and diseased groups, and might guide the design of studies yet to come, as it informs on a much larger fraction of the microbiome that might be changed.

Previous studies conducted on other individuals at high-risk of developing PD (e.g., isolated RBD-iRBD) also found that the gut microbiome of iRBD patients was at an intermediate state between healthy and early-stage PD individuals^25^. The body-first model of PD has gained attention in recent years, and results from our correlation analyses support the hypothesis that microbiome alterations might occur in the prodromal phase of body-first subtype of PD. Clinically, body-first PD patients are characterised by a higher burden of non-motor symptoms, in particular autonomic symptoms^26^. Our results showed that PD individuals with the most marked microbiome alterations had more severe non-motor symptoms. In non-parkinsonian individuals, we found similar results with autonomic dysfunction and depression being more severe in those people with more marked PD-like microbiome alterations. Remarkably, the 4 GBA-NMC individuals with the highest abundance of coherently PD-enriched species met the criteria for prodromal PD; this fits the expectation that 10-20% of *GBA1* variant carriers will develop overt disease (4 individuals out of n=43 included in our study) during their lifetime. These data highlight the importance of screening populations at-risk of PD (*GBA1* variant carriers, iRBD, etc.) based on a combination of clinical, genetic, and environmental factors, and then stratify them based on specific markers such as their gut microbiome profile. The lifetime risk for PD in the general population is approximately 3%^27,28^, therefore discovering PD-like microbiome alterations in ∼20% participants means that other genetic and/or environmental factors interact with and influence PD development.

In conclusion, we found that both genetically predisposed and healthy individuals without genetic risk can have gut microbiome changes like those typical of PD. As these alterations appear associated with disease progression in patients, and with the development of prodromal features in individuals at-risk of PD, screening of healthy individuals using a PDMS score such as the proof-of-concept PDMS-16, may enable identification of healthy individuals at higher risk of neurodegeneration. In such individuals, modification of factors such as diet, which is associated with microbiome alterations in our study, could help reduce the risk for PD^29^. Full assessment of the power of such approach to combat PD will have to await longitudinal follow-up of sufficiently large groups of individuals and for adequate length of time.

## Methods

### Study design

This was a case-control, multi-center, international study, with participants recruited from the United Kingdom and Italy. Four groups of individuals were enrolled: patients with Parkinson disease (PD) carrying a variant in the *GBA1* gene, patients with PD not carrying a variant in the *GBA1* gene, healthy controls (HC), and non-manifesting carriers of a *GBA1* variant (i.e., not diagnosed with PD, GBA-NMC). As healthy controls, we aimed to recruit PD patients’ partners to mitigate differences in diet and lifestyle habits and therefore the influence of these factors on the gut microbiome. Some HC individuals who were not partners/spouses of PD patients were also recruited as population control subjects.

### Recruitment strategy and *GBA1* gene sequencing

Recruitment strategy of study participants was performed, as previously described^1^. Participants from the UK were recruited via the RAPSODI GD and PD Frontline portals, from University College London (UCL)^2^. This study was approved by the local Ethics Committees (London – Queen Square REC: 15/LO/1155) and all participants signed informed consent upon enrolment. In the UK, the analysis of the *GBA1* gene was performed on saliva samples collected with the DNA OG-500 kit from DNA Genotek (https://www.dnagenotek.com/row/products/collection-human/oragene-dna/500-series/OG-500); after collection following the manufacturer’s instructions (https://www.dnagenotek.com/row/support/collection-instructions/oragene-dna/OG-500andOG-600), samples were posted back from study participants to UCL. DNA extraction and long range sequencing of an 8.9 kb amplicon, including all coding exons and introns of the *GBA1* gene, was performed using bespoke primers on the Oxford Nanopore MinION platform, as previously described^3^, partially at the laboratories of the Department of Clinical and Movement Neurosciences, UCL Institute of Neurology, United Kingdom, and partially at the Exeter Clinical Laboratory International, United Kingdom, a diagnostic clinical laboratory accredited by the UK’s National Accreditation Body (UKAS 8092). The *GBA1* positive results were confirmed by Sanger sequencing at the Exeter Clinical Laboratory International. Two healthy participants with synonymous variants (G264=/WT, and V499=/WT) have been considered as WT/WT for the *GBA1* gene, as synonymous variants are generally considered to be silent for any effect, since they do not change the amino acid sequence of the protein^4^. Participants from Italy were recruited from two neurology tertiary centres, i.e. the IRCCS Mondino Foundation in Pavia, and the Neurology Unit, Neuromotor & Rehabilitation Department, Azienda USL-IRCCS in Reggio Emilia. The sequencing of the *GBA1* gene in the Mondino Pavia cohort was performed by a next-generation sequencing (NGS)-based method, which includes the selective amplification of the whole *GBA1* gene in one long PCR fragment (6kb) followed by Nextera sequencing and a customized bioinformatics pipeline aimed at masking the *GBAP1* pseudogene. The *GBA1* positive results were confirmed by conventional Sanger sequencing through *GBA1* amplification in three overlapping fragments using specific primer pairs. Pathogenic variants in 15 PD-related genes linked to autosomal dominant (*SNCA*, *LRRK2*, *VPS35*, *GBA1*), X-linked (*RAB39B*), and autosomal recessive PD (*PRKN*, *PINK1*, *PARK7*, *ATP13A2*, *PLA2G6*, *DNAJC6*, *SYNJ1*, *FBXO7*, *VPS13C*, *PTRHD1*), were also evaluated using NGS-based sequencing of PD gene virtual panel as well as MLPA analysis (SALSA Kit P51-P52, MRC Holland). In the Reggio Emilia cohort, the sequencing of the *GBA1* gene for PD patients was performed by using a *GBA1*-specific long-range PCR with subsequent *GBA1* exon-specific PCR and NGS of the resulting products, as previously described^5^. PD patients were also tested for 11 pathogenic or likely pathogenic variants in the *LRRK2* gene, and if negative, 50 target genes were analysed by means of NGS-based sequencing of PD gene virtual panel^5^. For non-parkinsonian individuals, the genetic sequencing of *GBA1* was performed either by long PCR approach or NGS-based method^6,7^. This study was approved by the local Ethics Committees (EC of Pavia: code P-20210009687, and EC of Area Vasta Emilia Nord: code 2021/0092531), and written informed consent was obtained upon enrolment by all study participants.

### Clinical evaluation

Participants were invited for an in-person assessment in the clinic. Information about demographics and family history of PD was collected from each participant. Each participant underwent a clinical assessment to evaluate motor and non-motor symptoms of PD, including autonomic function, rapid eye movements-REM-sleep behaviour disorder, olfaction, mood, and cognition. The selected scales and questionnaires were the following: Movement Disorder Society-Unified Parkinson’s Disease Rating Scale MDS-UPDRS (part I-IV)^8^, Hoehn & Yahr (H&Y) scale^9^, SCales for Outcomes in PArkinson’s disease (SCOPA-AUT)^10^, Wexner Constipation Scoring System (WCSS)^11^, REM Sleep Behaviour Disorder Questionnaire (RBDSQ)^12^, University of Pennsylvania Smell Identification Test (UPSIT)^13^, Hospital Anxiety and Depression Scale (HADS)^14^, Beck Depression Inventory (BDI)^15^, and Montreal Cognitive Assessment (MOCA)^16^. For PD patients, information about time at diagnosis and medications was collected, and the levodopa equivalent daily dose (LEDD) was calculated^17,18^.

The MDS diagnostic criteria for prodromal PD were applied to the GBA-NMC group using the online Prodromal PD Calculator (https://www.movementdisorders.org/MDS/Members-Only/Prodromal-PD-Calculator.htm)^19^. Combining the estimated prior probability of PD, i.e., the age-adjusted prevalence of PD, with the total likelihood ratio (LR) of prodromal PD obtained by multiplying the LRs of each marker, a post-test probability percentage of developing PD can be obtained for each subject. In this study, the following markers were used: age, sex, coffee consumption, smoking status, *GBA1* genetic status, diagnosis of type II diabetes mellitus, physical inactivity, presence of RBD (as defined by screening questionnaire), history of excessive daytime somnolence, olfactory loss, constipation, urinary dysfunction, severe erectile dysfunction, depression, cognitive deficit, and subthreshold parkinsonism defined as MDS-UPDRS III total score above 6 (excluding postural and action tremor).

All participants provided information about their weight and height, and body mass index (BMI) was calculated. Participants were asked to provide information about their dietary habits, by completing a short form of food frequency questionnaire which has been previously validated against an extensive food quality questionnaire in a UK-based population^20^. A dietary quality score (DQS) based on fruit, vegetable, oily fish, non-milk extrinsic sugar (NMES) and fat intakes has been calculated using the DQS calculator provided by the authors.

Study data were collected and managed using REDCap electronic data capture tools hosted at University College London and IRCCS Mondino Foundation^21,22^.

### Sample collection

Participants were asked to collect at home a faecal sample, using the sample collection kit OMNIgene•GUT | OM-200 purchased from DNA Genotek, Ottawa, Ontario, Canada (https://www.dnagenotek.com/row/products/collection-microbiome/omnigene-gut/OM-200). Participants were instructed on how to collect and return the samples within maximum 7 days from collection (https://www.protocols.io/view/stool-sample-collection-protocol-261gerypjl47/v1). The OMNIgene•GUT | OM-200 stabilises microbial DNA from faeces. Returned samples were stored in freezer at -80°C until transfer to INRAE, MetaGenoPolis, Paris laboratories for analyses. Individuals with reported use of antibiotics during the month prior to collection of faecal samples were excluded from the metagenomics analysis.

### Statistical analysis for clinical data

Categorical variables were compared using χ2 test. Continuous variables are presented as means ± SDs, unless otherwise specified. Group demographics and continuous clinical variables were compared using t-test or non-parametric Wilcoxon rank-sum test for two-group comparison (HC vs GBA-NMC, and HC vs PD). For RBDSQ scores, data were categorised using clinically validated thresholds and ordinal logistic regression (OLR) was used to determine associations between categorised variables and the grouping variable, adjusted for covariates. MOCA scores were analysed using both a linear regression model and OLR model, adjusted for covariates. For UPSIT scores, a linear regression model was applied with adjustment for covariates. Statistical analyses of clinical data were performed using R version 4.2.3^23^.

### DNA extraction and high throughput sequencing

Frozen faecal materials were aliquoted to ≤ 1000µL with or without the addition of a liquefaction liquid (OMNIgene Liquefaction Reagent OM-LQR, DNAGenotek) to facilitate sample recovery. DNA extraction was performed following the procedure previously described in dx.doi.org/10.17504/protocols.io.dm6gpjm11gzp/v1, with the following modifications. The samples were transferred into a deep-well plate containing 400 µL of 0.1 mm glass beads (not in suspension) and centrifuged 3,486 ×g for twenty minutes prior to discarding the supernatant and adding 250 µL guanidium thiocyanate, 40 µL N-lauroyl sarcosine (10 % solution) and 500 µL N-lauroyl sarcosine (5 % solution in PBS 1X) to the samples. Subsequently, the sample plate was incubated at 70°C in a thermomixer for one hour, with stirring at 1,400 rpm. Following centrifugation of the plate at 3,486 ×g for a period of five minutes, the lysate was collected in a new plate. The pellet of the previous plate was then washed with 500 µL of TENP (50 mM Tris-HCL 20 mM EDTA 10 mM NaCl, saturated with PVPP). The plate was then vortexed and centrifuged at 3,486 ×g for five minutes, after which the recovered lysate was pooled with the previous one. Finally, the final lysate was centrifuged for a 10 minutes’ period at 3,486 ×g, after which 800 µL were collected in a new plate. This plate was employed for purification with magnetic beads on the QIASymphony. The utilised protocol has been designed for MGP with the QIAGEN DSP Virus/Pathogen kit. DNA was quantified using Qubit Fluorometric Quantitation (ThermoFisher Scientific, Waltham, US) and qualified using DNA size profiling on a Fragment Analyzer (Agilent Technologies, Santa Clara, US). Either 500ng or 1μg of high molecular weight DNA (>10 kbp) was used to build the library. Shearing of DNA into fragments of approximately 150 bp was performed using an ultrasonicator (Covaris, Woburn, US) and DNA fragment library construction was performed using the Ion Plus Fragment Library and Ion Xpress Barcode Adapters Kits (ThermoFisher Scientific, Waltham, US). Purified and amplified DNA fragment libraries were sequenced using the Ion Proton Sequencer (ThermoFisher Scientific, Waltham, US), with a minimum of 20 million high-quality 150 bp reads generated per library^24^.

### Read Mapping

We performed QC check to remove any low-quality sequences using Alientrimmer software v2.0^25^, with the following parameters: “-k 10 -l 45 -m 5 -p 40 -q 20”, and potential host-related reads with Bowtie2 v2.5.1^26^, and using the human reference genome Homo sapiens T2T-CHM13v2.0 (accession GCF_009914755.1)^27^. High-quality reads were mapped onto the 10.4 million gut human gene reference catalogue v1 (https://doi.org/10.15454/FLANUP)^28^, and the 8.4 million human oral microbial catalogue v1^29^, using the METEOR software v1^30^. Read mapping was performed in a two-step procedure, using an identity threshold of 95% to the reference gene catalogues. First, unique mapped reads were attributed to their corresponding genes. Second, shared reads were weighted according to the ratio of unique mapping counts. A downsizing procedure was performed to normalize gene counts between samples by randomly selecting a subset of reads depending on the sequencing depth (18 million reads for an average 24 million reads depth sequencing). The gene abundance table was then normalized using the FPKM strategy and analysed using MetaOMineR (momr) R package v1.31 (https://cran.r-project.org/web/packages/momr/index.html)^31^. Finally, we performed a final QC validation on the MSP species composition using the CroCoDeEL methodology (https://github.com/metagenopolis/CroCoDeEL)^32^, combined with a spearman correlations threshold of >0.65, to remove any low quality samples, and yielding the final cohort to 464 individuals (n=271 PD, n=150 HC and n=43 GBA-NMC individuals).

### MSP microbial species determination

The 10.4 million gut gene and the 8.4 million oral gene catalogues were previously organized into 1990 and 853 MSP species^29,33–35^, that correspond to clusters of co-abundant genes used as proxies for microbial species, and containing core and accessory genes. We removed duplicated species obtained from the catalogues, yielding to a total of 2055 MSPs, further filtered at a 10% occurrence threshold for a final MSPs species count at 627. Taxonomical annotation was assigned using the Genome Taxonomy Database GDTB R07-RS207^36^, using an in-house pipeline as described below. First, all genes were aligned on public databases (ncbi, wgs)^37^ using Blast^38^. MSP were annotated with the lowest taxonomical rank (from species to superkingdom) that brought consensus in at least 50% of its genes. To avoid misleading annotations due to error in databases, for each gene the 20 first hits were considered. MSP definition and taxonomy are available from Data INRAE (https://doi.org/10.15454/WQ4UTV and https://doi.org/10.15454/FLANUP). Relative abundance of a given MSP was computed as the mean abundance of its 100 ‘marker’ genes (that is, the genes that correlate the most altogether). If less than 10% of ‘marker’ genes were seen in a sample, the abundance of the MSP was set to 0. MSP richness was computed as the number of detected MSP species in a particular sample, before proceeding to occurrence filtering.

### Microbial functional potentials determination

To assess the functional potential of the gut microbiota at the module level, we used the METEOR software, which includes several functional databases as previously described^39^. Three databases were used to predict gene functions: Kyoto Encyclopedia of Genes and Genomes (KEGG) ^40^, eggNOG database (version 3.0)^41^, and TIGRFAM (version 15.0) ^42^. First, genes of the catalogues were annotated using KEGG107 database using Diamond^43^, and further clustered into functional pathway modules according to KEGG (Kyoto Encyclopaedia of Genes and Genomes) Orthology (KO) groups, Gut Metabolic Modules (GMM)^44^, and Gut Brain Modules (GBM)^45^. Second, KEGG, GMM and GBM modules were reconstructed in each MSP using their reaction pathways based on their detected annotated KO genes. GMM and GBM functional modules were selected because they are specific to gut bacterial and gut-brain axis functions. For each pair of MSP/individual, the completeness of any given functional modules was calculated by considering the set of genes detected in the MSP of each mouse and the MSP completeness in each mouse. For a given MSP in a specific individual, completeness of the modules was corrected by the abundance of the MSP. After correction, functional modules in each MSP/individual were considered as complete if at least 90% of the involved reactions were detected. Abundance of functional modules in each sample was computed as the sum of the MSP abundances containing the complete functional module.

### Computational analysis

Statistical analyses and figure computing were conducted using R environment V.4.4.1 (https://www.r-project.org/).

The comparison of species and functional modules between groups was performed using the testRelation2 function from the momr package V1.31^46^. This function generates a matrix that provides statistical results calculated with the Wilcoxon rank-sum test and quantifies the magnitude of these differences using Cliff’s Delta (CD) for each species and functional module.

Coherent and non-coherent enriched or depleted species and functional modules were identified based on the testRelation2 output for the HC vs PD and HC vs GBA-NMC comparisons, using CD direction as guide.

All linear models were performed using the lm function from the stats R package V.4.4.1^47^. Regression plots and barplots displaying CD values of coherent enriched and depleted species in the comparison HC vs PD and HC vs. GBA-NMC were created using the ggplot2 package V.3.5.1^48^, and equation formula was generated with the stat_poly_eq from the ggpmisc R package (V.0.6.0)^49^.

The contribution of coherent and non-coherent species (enriched and depleted) to the microbiome was evaluated by calculating their proportion and abundance for each sample. Proportion was computed as the number of cognate species present in the sample, divided by the total number of species in the sample. Abundance was computed as total abundance of cognate species divided by abundance of all species in the sample. Significance of the difference of abundance and prevalence between the study groups was estimated by the Student T-test performed with t_test function from rstatix R package (V.0.7.2)^50^.

Abundance and prevalence of cognate species in the quartiles of PD samples were computed by sorting the samples by a relevant parameter (abundance or prevalence of coherent or non-coherent, enriched or depleted species). The mean abundance and prevalence for the four quartiles and the average fold change of abundance and prevalence between the first and fourth quartiles were computed.

Abundance and prevalence of cognate species above and below the median in GBA-NMC samples were computed by sorting the samples by a relevant parameter (abundance or prevalence of coherent enriched species).

For each clinical variable, the significance of the difference between the study groups was assessed by the Student T-test with the t_test function.

Correlation between abundance of individual species or functional modules and clinical variables was computed by pairwise Spearman rank correlation tests, using cor.test function from stats R package. For each clinical variable significantly correlated with species and modules (pvalue < 0.05) the bias of PD-enriched and PD-depleted species was assessed by a chi-squared goodness-of-fit test. For each study group, barplots were created to display the number of species and functional modules showing positive and negative correlations with each clinical variable. The plots were generated using the ggplot2 package, with data manipulation facilitated by the dplyr package V.1.1.4^51^, tibble package V.3.2.1^52^, and tidyr package V.1.3.1^53^, and layout adjustments handled by the grid package V.4.4.1^54^. Finally, the barplots were combined into a single visualization using the ggarrange function from the ggpubr package V.0.6.0^55^, and wrap_plots function from the patchwork package V.1.2.0^56^.

Heatmaps illustrating the correlation values between species and clinical variables were generated using the Heatmap, Legend, and packLegend functions from the ComplexHeatmap package version 2.20^57^. The colour gradient was created using the colorRamp2 from the circlize R package V.0.4.16^58^. Pairwise Spearman rank correlation tests were performed using the rcorr function from the R package Hmisc version 5.1.3^59^.

Regression plots were created using the ggplot and stat_poly_eq functions. For each group, pairwise Spearman correlation tests were performed using the rcorr function from the Hmisc package, between the abundances of the 176 species that showed differences in the HC versus PD comparison and the clinical variables. Once correlation tables were generated for each study group, the tables for the PD and GBA-NMC groups were merged, as well as the tables for the HC and PD groups. For each merged table, a new variable was created to indicate whether the correlation values between each species and each clinical variable were in the same direction (positive or negative) between the two groups compared.

Other R packages were used for various tasks: to read data files in Excel format using the functions read_excel from readxl package (V.1.4.3)^60^, and read.xlsx from xlsx package (V.0.6.5)^61^, to manipulate variable names with the str_rename function from stringr package (V.1.5.1)^62^, to process data using the function foldchange from gtools package (V.3.9.5)^63^, dataframe format with the reshape2 package (V.1.4.4)^64^, and to create composite figures comprising multiple plots with the grid.arrange function from gridExtra package (V.2.3)^65^.

## Acknowledgment

This research was funded in part by Aligning Science Across Parkinson’s (Grant number: ASAP-000420) through the Michael J. Fox Foundation for Parkinson’s Research (MJFF) and by the EU Joint Programme—Neurodegenerative Research (JPND) through the MRC grant code MR/T046007/1. Additional funding was provided from the MetaGenoPolis grant ANR-11-DPBS-0001. For the purpose of open access, the author has applied a CC BY 4.0 public copyright license to all Author Accepted Manuscripts arising from this submission. RM and SY are supported by a Royal Free Charity fellowship.

## Author contributions

E.M., J.M., S.D.E., M.Al. and A.H.V.S. conceived and designed the study. E.M. and V.M. performed the clinical metadata processing and analysis. M.Av., C.G., R.M., S.Y., F.C., V.F., and G.T. contributed to the metadata processing. E.M., J.M., M.Av., C.G., P.M., L.G., S.L.D.P., R.M., S.K., N.Lo., S.Y., N.Li., F.V., F.C., V.F., and G.T. contributed to patient enrolment and data acquisition. M.Gi., A.F., C.M., A.D., B.Q. performed DNA extraction and metagenomic sequencing. V.M., Y.R., M.Ge, N.P., and M.Al. performed the metagenomic data pre-processing and quality control. E.M. performed and wrote codes for clinical data analyses. Y.R., M.Ge., V.M. and S.D.E. performed metagenomic data analyses. Y.R., M.Ge., V.M. and S.D.E. performed and wrote codes for data visualization. J.M., H.B., F.B., S.D.E., M.Al., and A.H.V.S. provided funding. E.M., S.D.E., and A.H.V.S. interpretated the results and wrote the first draft of the paper, with contribution from V.M., Y.R., M.Ge and M.Al. All the remaining authors edited and contributed to the final draft of the paper.

## Additional information

Supplementary information is available for this paper.

Correspondence and requests for materials should be addressed to Professor Anthony HV Schapira.

## Data availability statement

The clinical and metagenomics sequencing data that support the findings of this study will be deposited in a public data depository platform at the time of publication and available on request from the corresponding author, AHVS. The data are not publicly available due to restrictions e.g. their containing information that could compromise the privacy of research participants. The authors will provide access to data if required for peer review.

---

**Extended Fig. 1:**
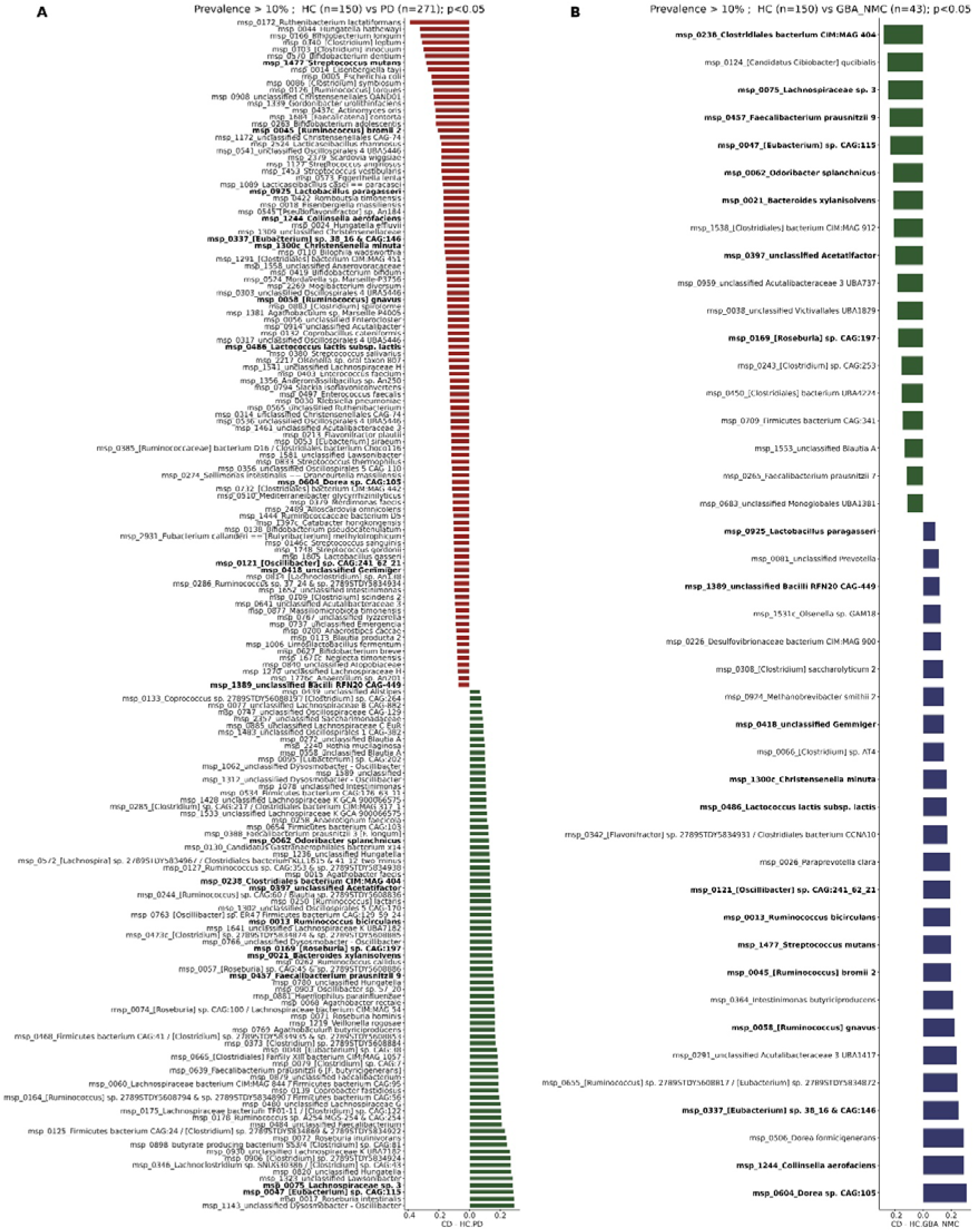
Gut microbiome variations in PD and GBA-NMC compared to HC. A. Barplot representing the 176 differentially abundant species observed in PD (n=271) compared to HC (n=150) individuals (Wilcoxon p<0.05). Species enriched and depleted in PD are shown in red and green, respectively. B. Barplot representing the 43 differentially abundant species observed in GBA-NMC (n=43) compared to HC (n=150) individuals (Wilcoxon p<0.05). Species enriched and depleted in GBA-NMC are shown in blue and green, respectively. The 21 significant species commonly altered between HC-PD and HC-GBA-NMC are highlighted in bold. All species except one (*msp_0013 Ruminococcus bicirculans*) are coherently altered between HC-PD and HC-GBA-NMC.

**Extended Fig. 2:**
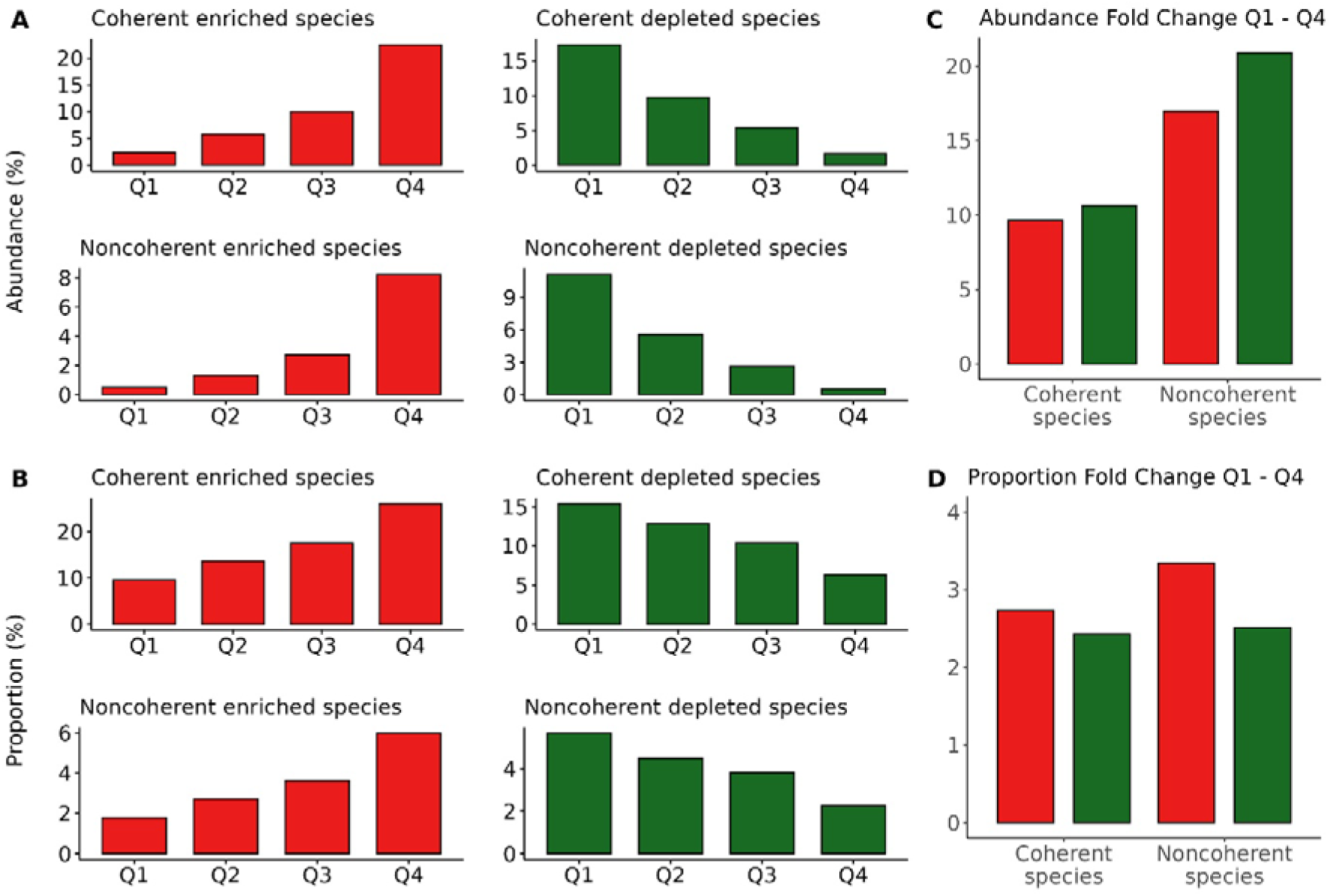
Microbiome variations in PD patients. A and B. Average values of abundance (A) and proportion (B) per participant (n=271) of the 176 significantly differentially abundant species found between PD and HC (PD enriched and coherent n=81, PD enriched and non-coherent n=18, PD depleted and coherent n=61, PD depleted and non-coherent species n=16) over the quartiles (Q) of individual distribution (Q1, 2 and 4; n=68, Q3; n=67 patients). C. Abundance fold change (FC) between Q1 and Q4. D. Proportion FC between Q1 and Q4.

**Extended Fig. 3:**
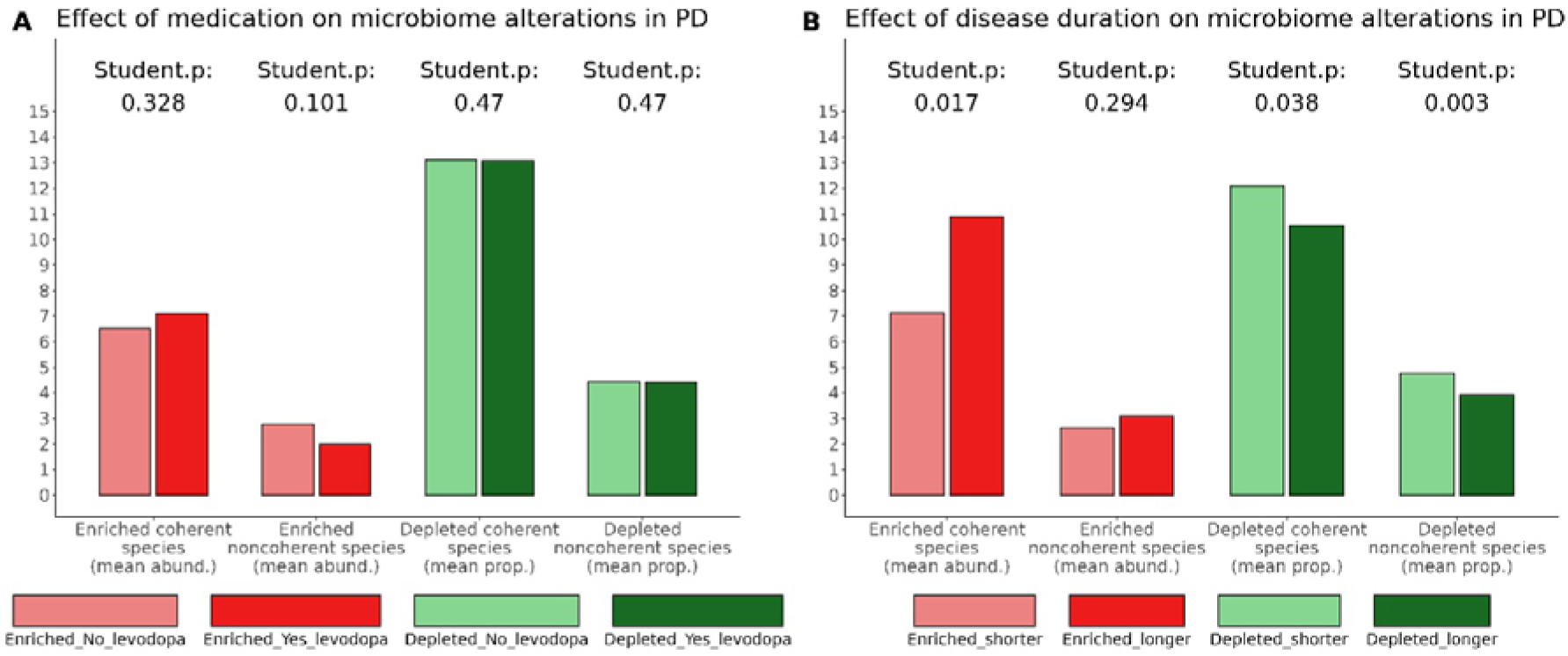
Effect of medication and disease duration on microbiome alterations in PD. A. Comparison of patients medicated (365 mgs, on average, n=47) with drug naïve (n=32), having on average similar disease duration (2.02 and 2.01 years, respectively). The highest possible number of medicated patients to match the average disease duration of all non-medicated patients of our cohort was included, to maximize statistical power of the analysis. B. Comparison of patients with different disease duration, taking, on average, similar medication levels (602 mgs on average). Shorter disease corresponds to 4.2 years, on average, (n=30) and longer to 7.2 years (n=31). P-value was computed by Student t test.

**Extended Fig. 4:**
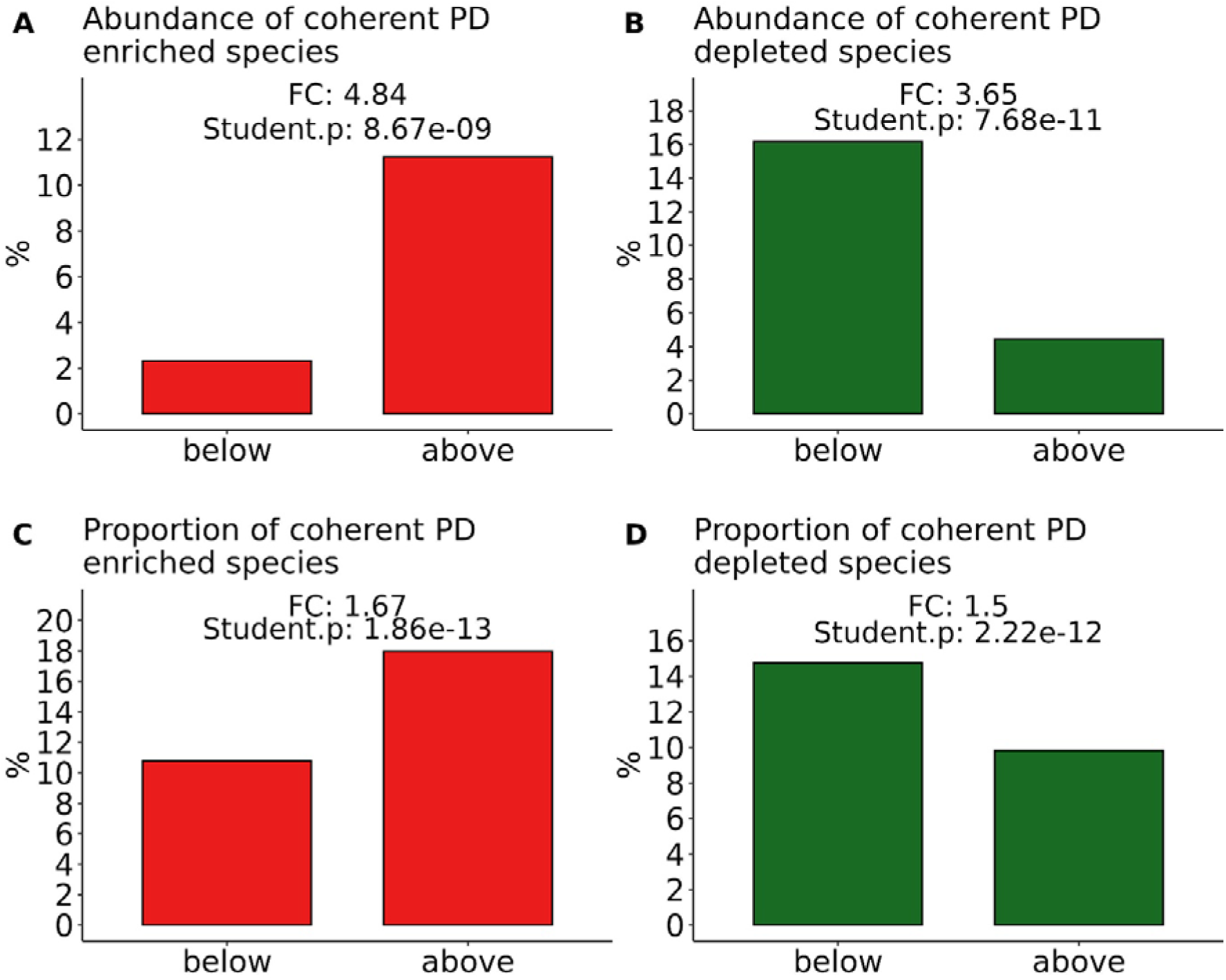
Microbiome variations in GBA-NMC. A and C. Average values of abundance (A) and proportion (C) of enriched coherent species above (n=22) and below (n=21) the median. B and D. Average values of abundance (B) and proportion (D) of depleted coherent species above (n=21) and below (n=22) the median. FC refers to Fold Change. P-value was computed by Student t test.

**Extended Fig. 5:**
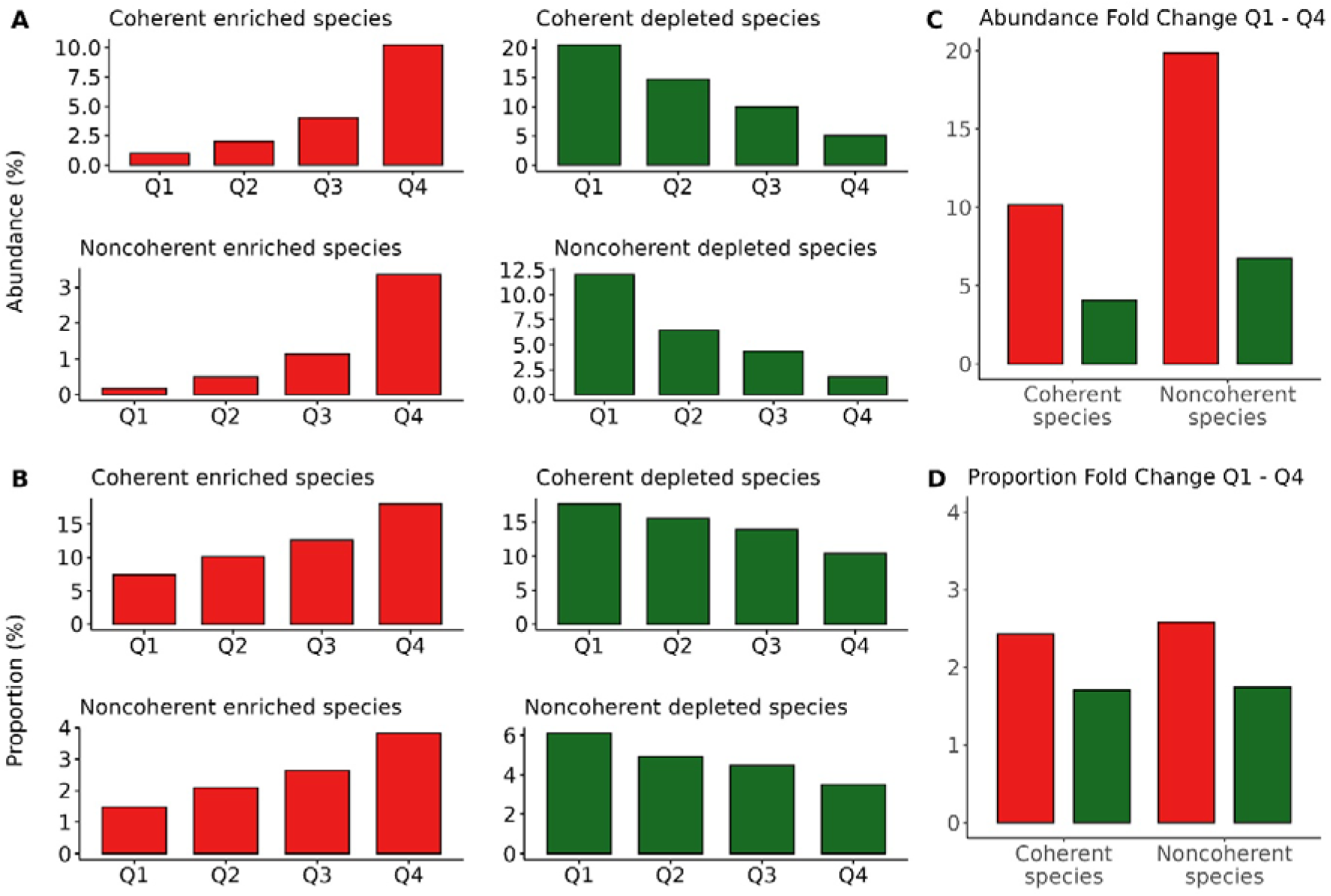
Microbiome variations in HC. A and B. Average values of abundance (A) and proportion (B) per participant (n=150) of the 176 significantly differentially abundant species found between PD and HC (PD enriched and coherent n=81, PD enriched and non-coherent n=18, PD depleted and coherent n=61, PD depleted and non-coherent species n=16) over the quartiles (Q) of individual distribution (Q1 and Q4; n=38, Q2 and Q3; n=37 individuals). C. Abundance fold change (FC) between Q1 and Q4. D. Proportion FC between Q1 and Q4.

**Extended Fig. 6:**
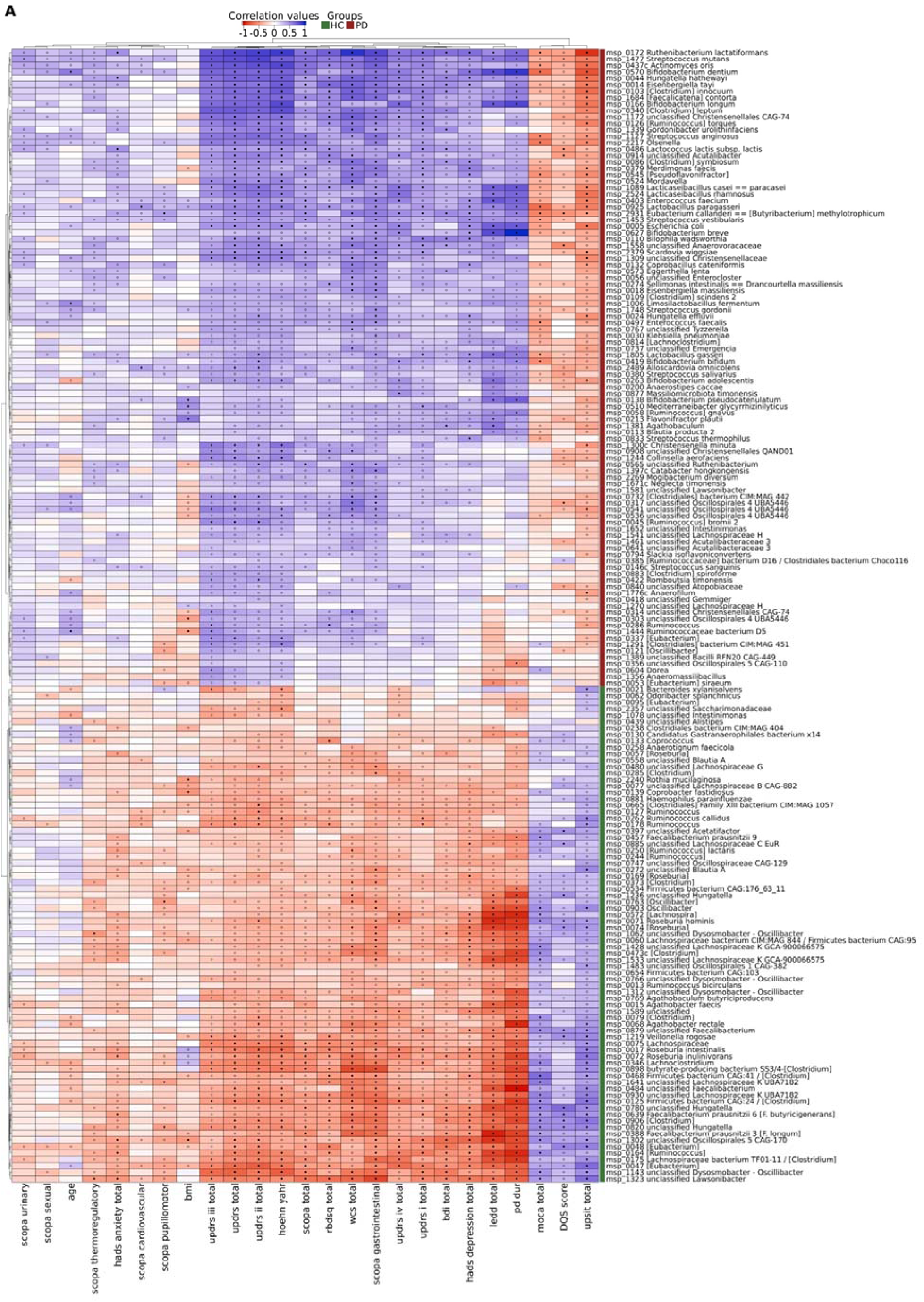
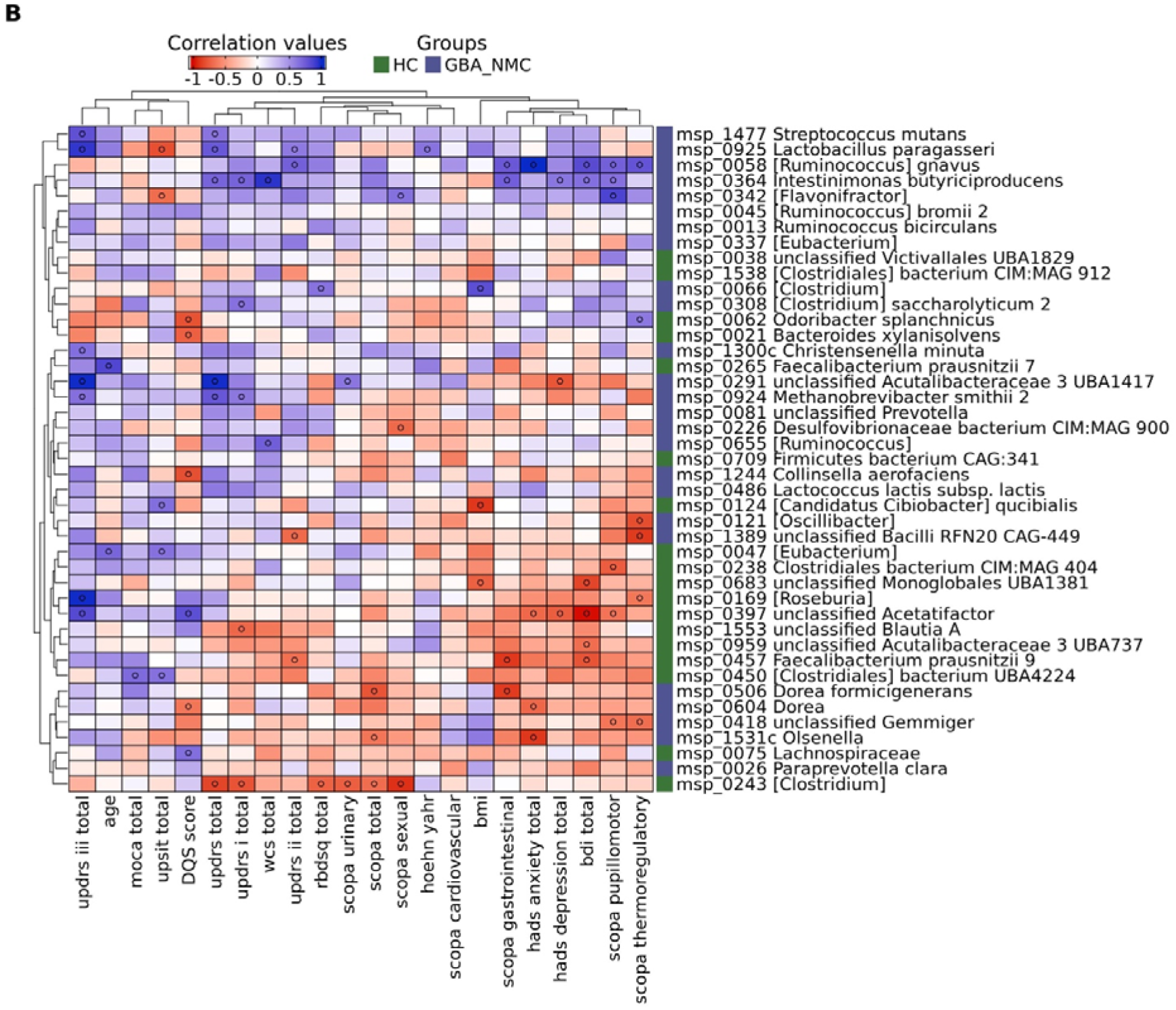
Heatmaps for Spearman correlations of differentially abundant species and clinical parameters. A. Correlation heatmap for the 176 differentially abundant MSP species enriched in HC (green) and PD (red) participants. B. Correlation heatmap for the 43 differentially abundant MSP species enriched in HC (green) and GBA-NMC (blue) participants. Positive correlations are indicated in blue, negative correlations in red. Open and solid dots referred to significant Spearman correlations of p<0.01 and p<0.05 respectively.

**Extended Fig. 7:**
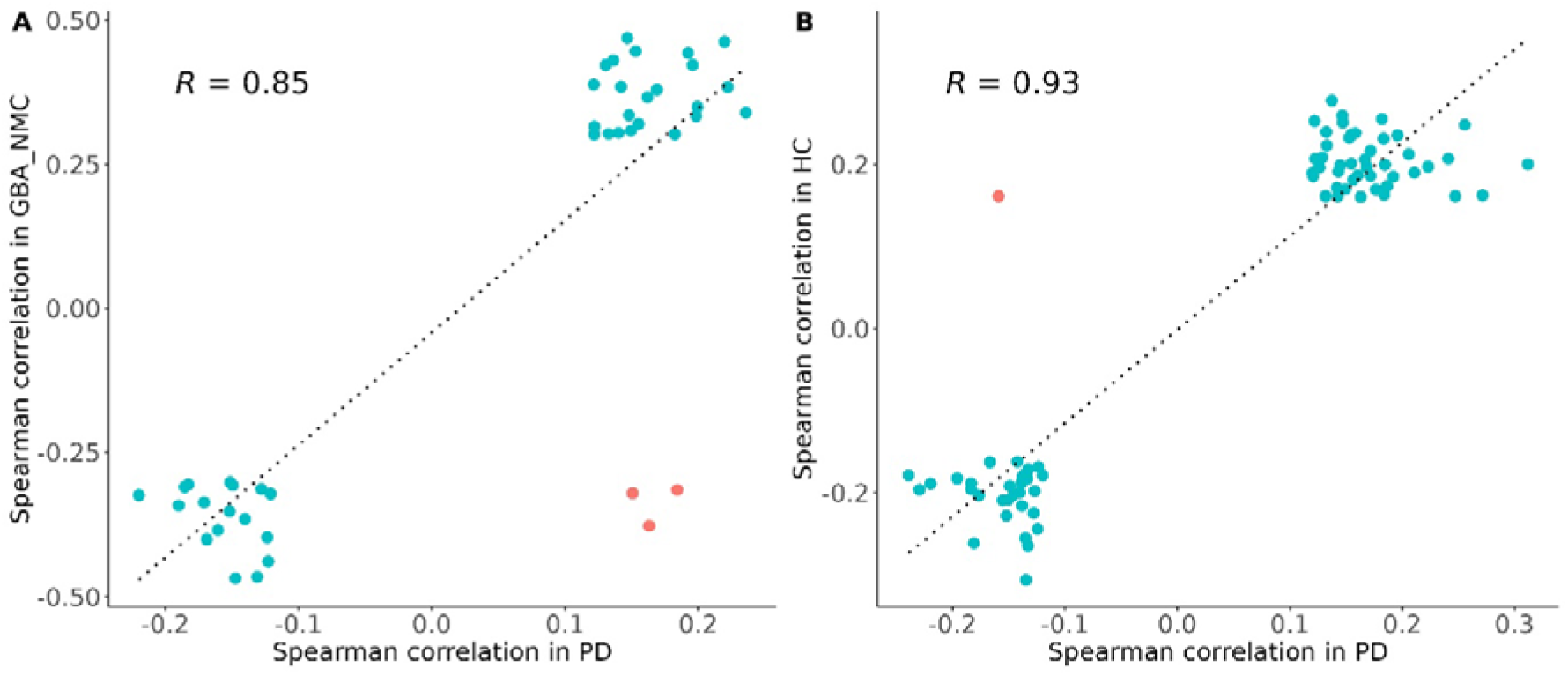
Significant Spearman correlation coefficients between species and clinical parameters in PD and GBA-NMC (A), or PD and HC (B). Blue dots indicate correlations with the same directions in both groups, and red dots indicate correlations with different directions in the two groups. R denotes the Pearson correlation coefficient between the Spearman values with the same direction.

**Extended Fig. 8:**
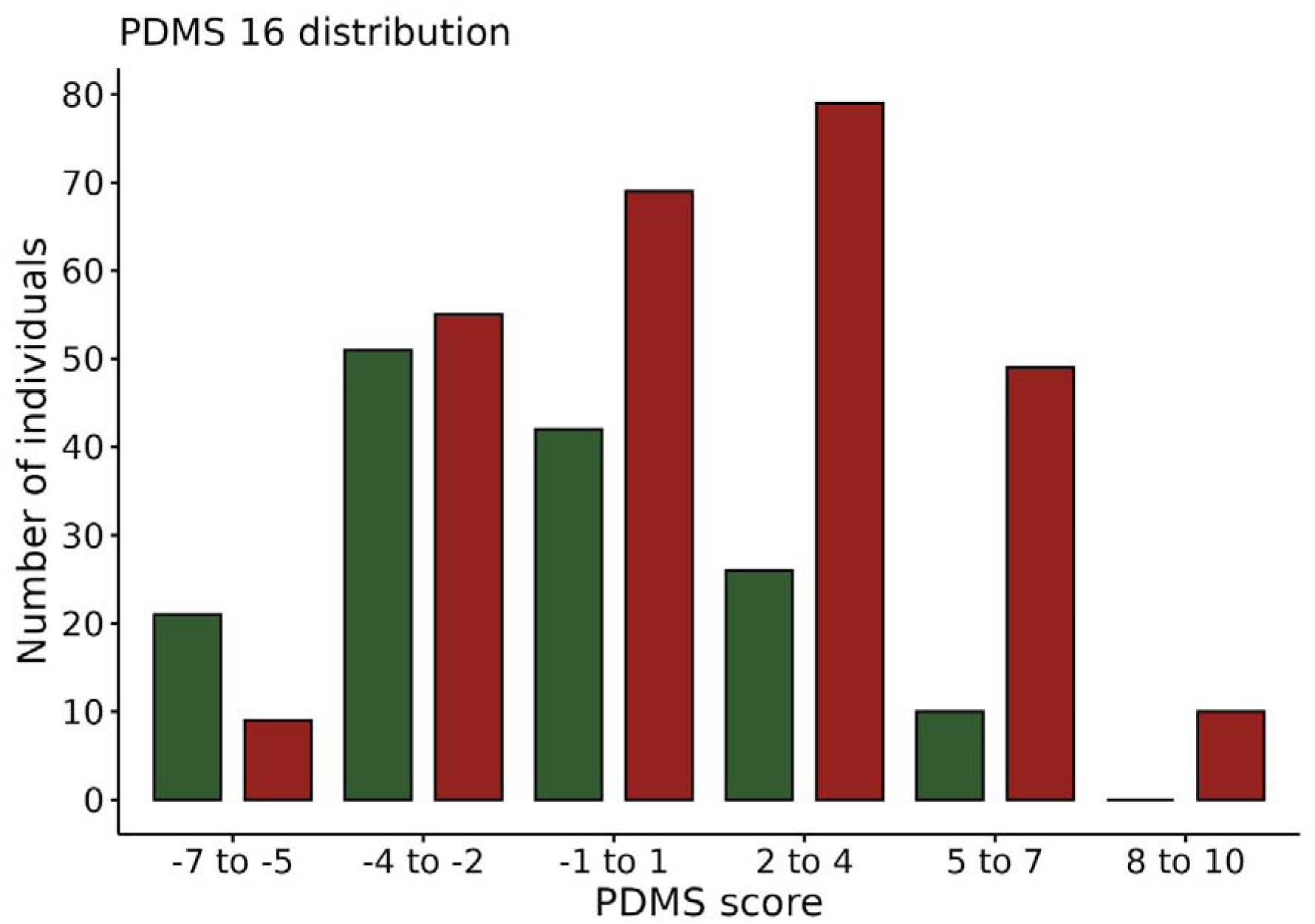
Distribution of PDMS-16 in HC (n=150) and PD (n=271) groups. Green and red bars refer to HC and PD, respectively.

**Extended Fig. 9:**
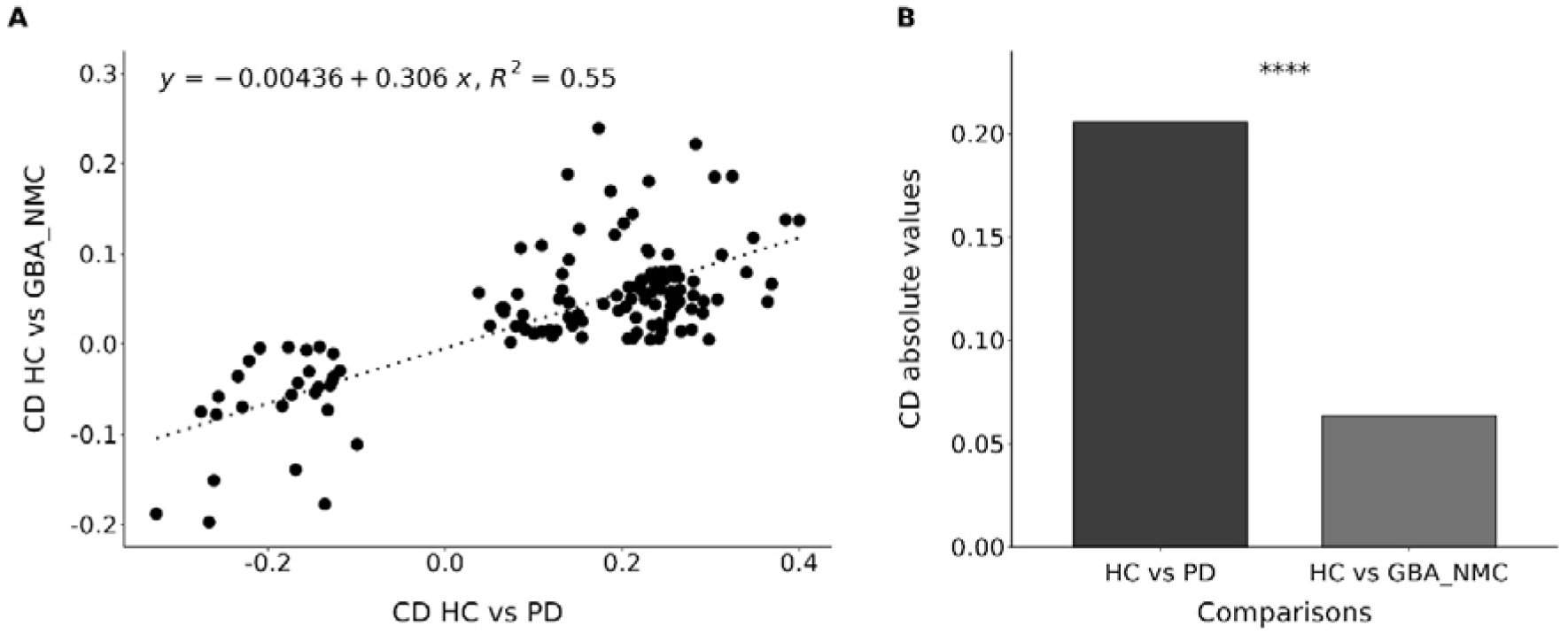
Variations of functional modules in PD and GBA-NMC relative to HC. **A.** Correlation of Cliff Delta for coherently varying modules, either enriched or depleted. R^2^ indicates correlation coefficient squared. **B**. Absolute values of Cliff delta in HC vs PD or GBA-NMC. P value was computed by the Student test; **** for p<0.0001, *** for p< 0.001, ** for p<0.01 and * for p<0.05.

**Extended Table 1.** Association of clinical variables and microbiome alterations. PD patients. Comparison of top (n=68) and bottom (n=68) quartiles; the values shown were obtained with the abundance of coherent depleted species, the number of statistically significant tests (enriched and depleted coherent and non-coherent species, by abundance and proportion) is shown. Variables significant in 6 or more tests are highlighted in bold. GBA-NMC individuals. Comparison of individuals below and above the median; the significant values were obtained using the proportion of coherently enriched/depleted species, bold print indicates significant values by both tests. HC individuals, proportion of species. Comparison of individuals in the top (n=38) and bottom (n=38) quartiles; the values shown were obtained with the proportion of coherent PD-enriched species. In bold are the variables that were also significant with the proportion of PD depleted species, significant difference of the UPDRS part I score was found in the latter but not the former comparison (in italics). HC individuals, PDMS-16. Comparison of individuals having PDMS-16 of -5 or less (lower microbiome alteration, n=21) with those having PDMS-16 of 3 or more (higher microbiome alteration, n=27); significantly different parameters in comparison of the latter individuals with the remainder of the HC cohort (n=123) are indicated by p-values in bold print, significant difference of the RBDSQ score was found in the latter but not the former comparison (in italics). Legend: n, no; y, yes; f, female; m, male; BMI, body mass index (m/kg²); BDI, Beck Depression Inventory; DQS, Dietary Quality Score; HADS, Hospital Anxiety and Depression Scale; UPDRS, Unified Parkinson’s Disease Rating Scale; MOCA, Montreal Cognitive Assessment; PD, Parkinson disease; SCOPA, SCales for Outcomes in PArkinson’s disease; RBDSQ, REM Sleep Behavior Disorder Questionnaire; UPSIT, University of Pennsylvania Smell Identification Test; WCSS, Wexner Constipation Scoring System.

| Microbiome alteration | PD |  |  |  | GBA-NMC |  |  | HC, proportion of species |  |  | HC, PDMS-16 |  |  |
| --- | --- | --- | --- | --- | --- | --- | --- | --- | --- | --- | --- | --- | --- |
|  | Low er | Hig her | t-test p | test p<0.05 | Low er | Hig her | t-test p | Low er | Hig her | t-test p | Low er | Hig her | t-test p |
| PD duration, years | 4,6 | 9,4 | 2,16E-09 | 8 |  |  |  |  |  |  |  |  |  |
| Levodopa dose, mgs | 423 | 806 | 4,53E-07 | 8 |  |  |  |  |  |  |  |  |  |
| BDI score | 9,1 | 11,0 | 4,53E-02 | 7 | 4,0 | 7,0 | 3,26E-02 | 3,5 | 5,9 | 7,56E-03 | 3,5 | 6,0 | 8,89E-03 |
| HADS depression score | 4,6 | 6,2 | 6,47E-03 | 7 | 1,8 | 3,2 | 3,27E-02 | 1,6 | 3,2 | 6,88E-03 | 1,2 | 3,3 | 1,59E-03 |
| HADS anxiety score | 5,4 | 6,3 | 1,03E-01 | 1 | 3,2 | 5,3 | 2,40E-02 | 2,8 | 4,3 | 1,39E-02 | 2,5 | 4,9 | 3,17E-03 |
| MDS-UPDRS I score | 9,1 | 11,6 | 1,28E-02 | 3 | 3,7 | 6,5 | 1,48E-02 | 2,5 | 3,9 | 4,04E-02 | 2,1 | 4,7 | 1,34E-02 |
| MDS-UPDRS II score | 9,2 | 12,3 | 4,10E-03 | 6 | 0,3 | 1,8 | 2,75E-03 | 0,1 | 0,6 | 3,03E-03 |  |  |  |
| MDS-UPDRS III score | 25,0 | 27,6 | 1,11E-01 | 1 |  |  |  |  |  |  |  |  |  |
| MDS-UPDRS IV score | 2,1 | 3,8 | 1,14E-03 | 8 |  |  |  |  |  |  |  |  |  |
| MDS-UPDRS total score | 45,4 | 55,3 | 4,59E-03 | 5 | 7,5 | 11,3 | 4,17E-02 | 4,4 | 6,6 | 3,89E-02 | 4,4 | 7,6 | 3,19E-02 |
| Hoehn and Yahr scale | 1,8 | 1,9 | 1,89E-01 | 5 |  |  |  |  |  |  |  |  |  |
| SCOPA gastrointestinal score | 3,3 | 4,8 | 2,19E-03 | 8 | 0,8 | 1,7 | 8,44E-03 | 0,8 | 1,4 | 2,73E-02 | 0,4 | 2,0 | 2,01E-05 |
| SCOPA pupillomotor score | 0,4 | 0,6 | 6,61E-02 | 4 | 0,2 | 0,7 | 9,92E-03 | 0,2 | 0,4 | 3,38E-02 | 0,3 | 0,8 | 2,04E-02 |
| SCOPA cardiovascular score | 0,8 | 0,8 | 4,38E-01 | 0 |  |  |  |  |  |  |  |  |  |
| SCOPA thermoregulatory score | 1,9 | 2,6 | 4,49E-02 | 1 | 0,8 | 1,8 | 2,01E-02 | 0,7 | 1,4 | 9,13E-03 | 0,7 | 1,6 | 1,67E-02 |
| SCOPA urinary score | 4,7 | 4,6 | 4,53E-01 | 0 |  |  |  |  |  |  |  |  |  |
| SCOPA sexual score | 1,8 | 1,5 | 1,89E-01 | 0 |  |  |  | 0,6 | 1,2 | 1,44E-02 | 0,7 | 1,4 | 2,03E-02 |
| SCOPA total score | 12,8 | 15,3 | 2,69E-02 | 4 | 6,4 | 9,1 | 4,25E-02 | 6,0 | 8,1 | 3,02E-02 | 4,8 | 9,9 | 8,62E-04 |
| RBDSQ | 4,9 | 6,0 | 4,93E-02 | 4 | 1,3 | 3,2 | 3,95E-03 |  |  |  | 1,52 | 2,26 | 4,36E-02 |
| Sleepiness | 0,9 | 1,3 | 9,85E-03 | 5 |  |  |  | 0,3 | 0,6 | 4,73E-02 |  |  |  |
| WCSS score | 4,9 | 7,9 | 5,46E-05 | 8 |  |  |  | 1,8 | 3,1 | 8,67E-03 | 1,9 | 4,1 | 1,42E-03 |
| Appendectomy: n=1, y=2 | 1,2 | 1,4 | 8,18E-03 | 7 |  |  |  | 1,1 | 1,3 | 2,23E-03 | 1,1 | 1,4 | 3,76E-03 |
| Appendectomy prior to PD, years | 43,2 | 35,6 | 4,90E-02 | 6 |  |  |  |  |  |  |  |  |  |
| Age, years | 62,8 | 63,6 | 3,10E-01 | 1 | 62,0 | 60,4 | 3,13E-01 | 61,3 | 62,6 | 2,86E-01 | 62,9 | 60,1 | 1,92E-01 |
| BMI | 25,4 | 25,5 | 4,28E-01 | 1 | 25,5 | 26,3 | 2,63E-01 | 25,1 | 26,5 | 9,61E-02 | 25,5 | 25,6 | 4,71E-01 |
| Sex: f=1, m=2 | 1,6 | 1,5 | 8,59E-02 | 3 | 1,5 | 1,4 | 3,33E-01 | 1,4 | 1,4 | 2,45E-01 | 1,4 | 1,3 | 1,76E-01 |
| UPSIT total | 19,6 | 16,7 | 2,29E-03 | 7 |  |  |  |  |  |  |  |  |  |
| MOCA score | 27,2 | 25,0 | 6,28E-05 | 8 |  |  |  |  |  |  |  |  |  |
| DQS score | 11,3 | 10,3 | 2,14E-04 | 8 |  |  |  | 11,3 | 10,7 | 4,51E-02 | 11,9 | 10,7 | 1,14E-03 |
| Fruit, portions | 2,2 | 1,9 | 6,86E-04 | 7 | 2,1 | 1,9 | 2,96E-02 | 2,3 | 2,0 | 1,14E-02 | 2,3 | 2,0 | 2,22E-02 |
| Vegetables, portions | 2,0 | 1,8 | 3,81E-02 | 6 |  |  |  | 2,1 | 1,8 | 1,49E-02 | 2,3 | 1,6 | 6,77E-05 |
| Education, years | 15,8 | 13,0 | 2,46E-05 | 8 |  |  |  | 16,7 | 13,4 | 3,74E-04 | 19,0 | 14,7 | 1,26E-03 |

